# More severe pneumonitis in children predicts the need for admission and elevation of some but not all markers of severe Covid-19

**DOI:** 10.1101/2022.03.19.22272644

**Authors:** Paul Walsh, Andrea Hankins, Heejung Bang

## Abstract

Unlike most other viral pneumonitis, SARS-CoV-2 often causes hyperferritinemia, elevations in D-dimer, lactate dehydrogenase (LDH), transaminases, troponin, CRP, and other inflammatory markers. We questioned (1) if the severity of pneumonitis observed on lung ultrasound was associated with hospitalization and (2) could lung ultrasound be used to stratify which children needed blood tests?

**Methods:** We did a retrospective cross-sectional review of children aged between 14 days and 21 years of age being evaluated for Covid-19 in our pediatric emergency department from 30/November/2019 to 14/August/2021 who had had a point-of-care lung ultrasound. Lung ultrasounds were categorized using a 6-point ordinal scale. We used logistic regression to estimate the adjusted effect of lung ultrasound on hospital admission. We performed ordinary least square regression for the association between lung ultrasound severity and laboratory abnormalities. We adjusted these using propensity score derived inverse probability weighting to account for the non-random decision to obtain laboratory investigations.

**Results:** We identified 500 point-of-care lung ultrasounds of which 427 could be assigned a severity category. Increasing lung ultrasound severity was associated with increased hospital admission OR 1.36(95% CI 1.08, 1.72.) Ferritin, LDH, transaminases, and D-dimer, but not CRP or troponin were significantly associated with more than moderately severe lung ultrasounds. D-Dimer, CRP, and troponin were sometimes elevated even when lung ultrasound was normal.

**Conclusion:** Severity of pneumonitis was associated with hospital admission. Ferritin, LDH, transaminases, and D-dimer were increased in more than moderately severe pneumonitis but lung ultrasound did not predict elevation of other markers.

## Introduction

Bedside lung ultrasound allows the immediate diagnosis of viral pneumonitis in children who present with a viral respiratory infection. Lung ultrasound enhances diagnostic precision; if there is no pneumonitis and nasal or nasopharyngeal nucleic acid testing is negative SARSCoV-2 and the diagnosis of covid-19 has effectively been excluded. Lung ultrasound has been validated for the diagnosis of covid pneumonitis using a CT gold standard in adults and older children[1] and has also been shown to predict which patients are more likely to progress.[2,3] Certain blood tests have also been shown to herald increased morbidity or mortality e.g. Ferritin [4–7] LDH [8–10], ALT, AST, [8,11] D-dimer[12,13], CRP[8], ESR[8], Procalcitonin[8], BNP[8], and possibly WBC[8]. Others, such as troponin elevations are considered a *sine qua non* for the diagnosis of myocarditis.

This raises two questions: First, does the severity of lung ultrasound abnormalities predict hospitalization? If lung ultrasound fails this test of validity it is of little use. Second, if the lung ultrasound is normal can the clinician confidently skip blood testing? And how abnormal would a lung ultrasound have to be to allow us to avoid blood tests? Such an approach is inherently attractive in the pediatric emergency department (ED); bedside lung ultrasound can be performed rapidly and is minimally invasive. Conversely, obtaining blood work in children imposes substantial costs in terms of staff time and distress to both the child and parents.

Answering these questions is hindered by selection bias. The decision to obtain blood tests on a patient may also be influenced by the ultrasound findings. This leads to potential incorporation bias if not correctly analyzed. Here we are sidestepping the issues related to the decision to obtain point-of-care lung ultrasound and are limiting our questions to those in whom lung ultrasound was performed. We address potential selection bias for obtaining laboratory studies with propensity score analysis.

## Methods

This was a retrospective cross-sectional chart review. Patients were eligible if they were being evaluated for possible SARS-CoV-2 infection, were aged between 14 days and 21 years of age, were seen in our ED, and had a point of care lung ultrasound reported in the chart. Data collection for this study includes data from 30/November/2019 to 14/ August/2021. Chart selection and extraction were performed electronically using regexm functions for SARS-CoV-2 and Covid rather than ICD-10 codes. We did this because ICD codes applied at discharge from clinical settings do not necessarily capture the clinical diagnosis.[14]

### Data sources

All data was obtained from the electronic medical record. Laboratory data were extracted from the electronic medical record. Reference laboratory values were taken from the reference ranges provided by the laboratory for each individual patient. For procalcitonin, we took a level of > 0.50 ng/mL as abnormal. Only laboratory values obtained during the emergency department (ED) visit blood draw were included. The SQL code used to extract laboratory data is in the Appendix.

### Ultrasound

One author (PW) reviewed all the lung ultrasounds using a template to assign severity. Ultrasound reports were extracted as text snippets and presented to the reviewer without access to the remainder of the patient chart. The reviewer categorized each ultrasound using a 7-point scale: normal, or as having very mild, mild, mild to moderate, moderate, moderate to severe, or severe pneumonitis. Because of small numbers in the ‘moderate to severe’ and ‘severe’ categories, these were combined into one category in the analysis phase. Ultrasound reports rather than raw images were used because (1) in community practice bedside point-of-care lung ultrasounds frequently include only representative images rather than clips of every window (2) the written ultrasound report allows us to know how the physician treating the patient interpreted the findings. The ultrasound reviewer was not blinded to the study questions.

### Statistical analysis

#### Intra-rater reliability of lung ultrasound report interpretation

Each report was presented multiple times in a slightly different text fragment without access to the rest of the medical record. Where different determinations of lung ultrasound severity were made the more severe category was used in the analysis.

A subset of 71 patients had their final ultrasound report re-evaluated by the original reviewer blinded to the original report. This re-evaluation occurred more than one month after the original evaluations were performed. The intra-rater agreement was measured using Gwet’s AC-1 coefficient and intraclass correlation coefficient (random-effects model). These are preferred to @ statistics when the agreement is expected to be high. [15] [16,17]. Intra-rater reliability was measured using the original seven classifications of disease severity.

#### The relationship between lung ultrasound findings and hospital admission

We performed the Cochran-Armitage test for a trend of the severity of lung ultrasound findings and hospitalization. We also performed this test for oxygen saturation (measured at triage) and the initial ESI triage category. Oxygen saturation was modeled both using a dichotomous threshold of 92%, using oxygen saturation grouped as follows, 97%-100%; 94%-96%; 92%-93%; 91%-90%, <90%, and <85%, and as a continuous variable. We included ultrasound severity, oxygen saturation, and the emergency severity index (ESI)[18] triage category in multivariable-adjusted analysis using logistic regression. Ultrasound severity (0-6) and ESI triage category (0-5) were modelled as continuous variables. We observed multicollinearity between the ESI triage category and oxygen saturation. Models with ESI category and ultrasound severity had better fit characteristics than those with oxygen saturation or both ESI category and oxygen saturation and were reported.

#### The relationship between lung ultrasound and laboratory findings

The decision to obtain laboratory investigations in these children suspected of having Covid-19 is non-random and almost always made before the results from nasal PCR swabs can be available. We initially performed ANOVA tests with the value of the blood test as the dependent and lung ultrasound severity as the independent variable. We also performed logistic regression dichotomizing lung ultrasound severity category as being above or below the highest two levels of severity. We performed the Jonckheere–Terpstra test for a trend for each lab test result for ultrasound severity. Any blood test with a p-value < 0.10 in both the ANOVA and logistic regression was further analyzed by propensity score analysis.[19,20]

We estimated inverse probability weights for each of these blood tests using logistic regression-based propensity analysis scores. These scores included the ultrasound findings; ESI triage category; and for some blood tests admission; with the precise specification depending on fit characteristics of the model. These inverse probability weights were then used to weight ordinary least squares regression analysis. This was done to account for the non-random decision to obtain blood tests which were likely influenced by the overall severity of illness of the child (reflected in the ESI category) and the severity of the lung ultrasound findings. Logistic regression was used because the decision to obtain blood work is ultimately binary, even if the factors leading to that decision are not.

In the case of troponin and D-dimer, both assays were changed during the study period and no calibration between the two tests was provided. Therefore, we dichotomized each at the 99th centile and analyzed these as normal or abnormal using logistic regression. In the case of D-dimer, dichotomizing lung ultrasound severity category as being moderately severe was necessary to avoid perfect prediction of elevated d-dimer in the presence of ‘more than moderately severe’ lung ultrasound findings. An alternative analysis using exact logistic regression which allows for this scenario is in the appendices. We used Stata17.0 (Statacorp LLP, College Station, TX) for data analyses.

### Sensitivity analysis

In sensitivity analysis, we repeated our final models for each significant laboratory test but included age group as a dummied variable using school-age children as the referent category. We did this because of the broad range of ages of our patients. The age groups we used were: Neonate 14-28 days, infant, 29 days to one year; preschooler 1 to 5 years; school-age 6 to 12 years, teenager, 13 to 18 years; transitional, 18 to 21 years. We excluded neonates younger than 14 days because of the potential for residual pulmonary hypertension making lung ultrasound interpretation more difficult. Our PED sees all children covered by California Children’s Service (CCS) up to age 21 years of age, and depending on capacity may also see non-CCS-covered children up to 21 years of age. We analyzed age group rather than age because age is likely nonlinear and these categories, although uneven, better express childhood. These analyses could answer the question: Are there differences between age groups in these outcomes?

## Results

There were 500 point-of-care lung ultrasounds mentioned in patient medical records of which 427 (each representing a distinct visit in 371 distinct patients) could be assigned a severity category based on the physicians’ written ultrasound report. The median patient age was 6 years and 229/427 (54%) were male. Patient characteristics are shown in **Table 1**. Thirty-eight, (9%) patients were admitted and 84 (20%) had at least one blood test performed. Nasal turbinate or nasopharyngeal testing was performed on 398 visits and was positive in 108 (27%). Another virus was identified in 98 visits, most commonly rhinovirus (73), another coronavirus (10), or a parainfluenza virus (7).

**Table 1.** Patient characteristics. ED emergency department; SD standard deviation; IQR interquartile range; ESI emergency severity index.

| Factor | Level/Unit | Value |
| --- | --- | --- |
| Sample size, N |  | 427 |
| Male |  | 229 (54%) |
| Age, mean +/- SD | (years) | 8.1 +/- 6.9 |
| Age, median (IQR) | (years) | 6.2 (1.7, 13.9) |
| Age by category | Neonate < 1 month | 10 (2%) |
|  | Infant 1 to 12 month | 58 (14%) |
|  | Preschooler 1-5 years | 140 (33%) |
|  | School-age 6-12 years | 88 (21%) |
|  | Teenager 13-18 | 84 (20%) |
|  | Transitional 18 to 21 years | 47 (11%) |
| ESI triage category | 1 | 1 (0.2%) |
|  | 2 | 37 (9%) |
|  | 3 | 116 (27%) |
|  | 4 | 255 (60%) |
|  | 5 | 18 (4%) |
| ED length of stay median (IQR) | (minutes) | 122 (83, 190) |
| ED length of stay if discharged | (minutes) | 118 (80, 171) |
| Blood test obtained | Any/None | 84 (20%) |
| Admitted |  | 38 (9%) |

Intra-rater agreement for the classification of ultrasound reports was high; Gwet’s AC-1 coefficient for agreement was 0.82, and the intra-class correlation for intra-rater reliability for lung ultrasound was 0.92.

Most lung ultrasounds were normal or showed only mild disease. Lung ultrasound findings are summarized in **Table 2**.

**Table 2.**
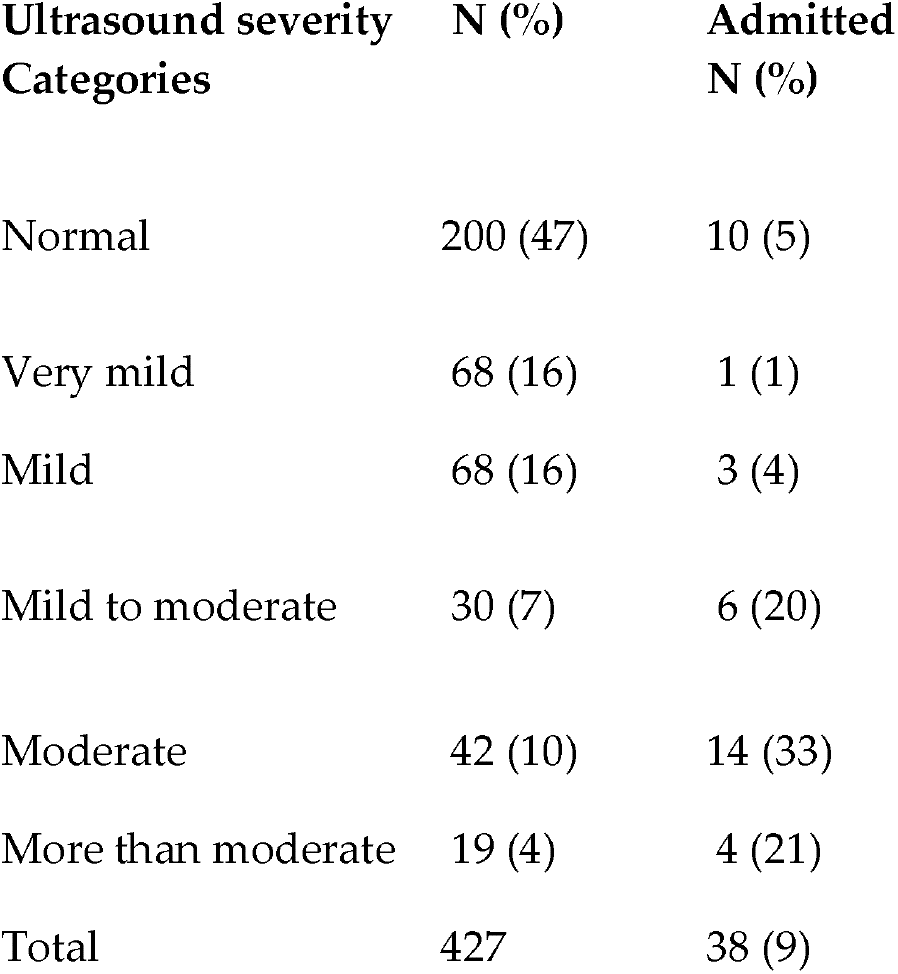
Lung ultrasounds’ severity originally recorded in the medical record was grouped for analysis; the ‘More than moderate’ category includes 7 moderate to severe and 12 severe cases. Percentages in parentheses. Percentages are affected by rounding.

| Ultrasound severity Categories | N (%) | Admitted N (%) |
| --- | --- | --- |
| Normal | 200 (47) | 10 (5) |
| Very mild | 68 (16) | 1 (1) |
| Mild | 68 (16) | 3 (4) |
| Mild to moderate | 30 (7) | 6 (20) |
| Moderate | 42 (10) | 14 (33) |
| More than moderate | 19 (4) | 4 (21) |
| Total | 427 | 38 (9) |

### Admission rates

Admission tended to increase with increasing severity of lung ultrasound (**Table 2**) The Cochrane-Armitage test for trend was significant (*p* <0.000) but with evidence of departure from a linear trend (Pearson). Admission rates showed similar trends by triage categories (*p*<0.0001) with evidence of non-linearity, and with oxygen saturation grouped at clinically meaningful levels (*p*=0.0012). In multivariable adjusted regression lung ultrasound severity OR 1.36(95% CI 1.08, 1.72,) and ESI triage category OR (0.11 95% CI 0.06, 0.20) using ESI category 1 as referent, were significant predictors of admission. **Figure 1** shows the probability of admission based on lung ultrasound findings at different ESI levels. Lung ultrasound findings are only part of the explanation of hospital admission.

**Figure 1.**
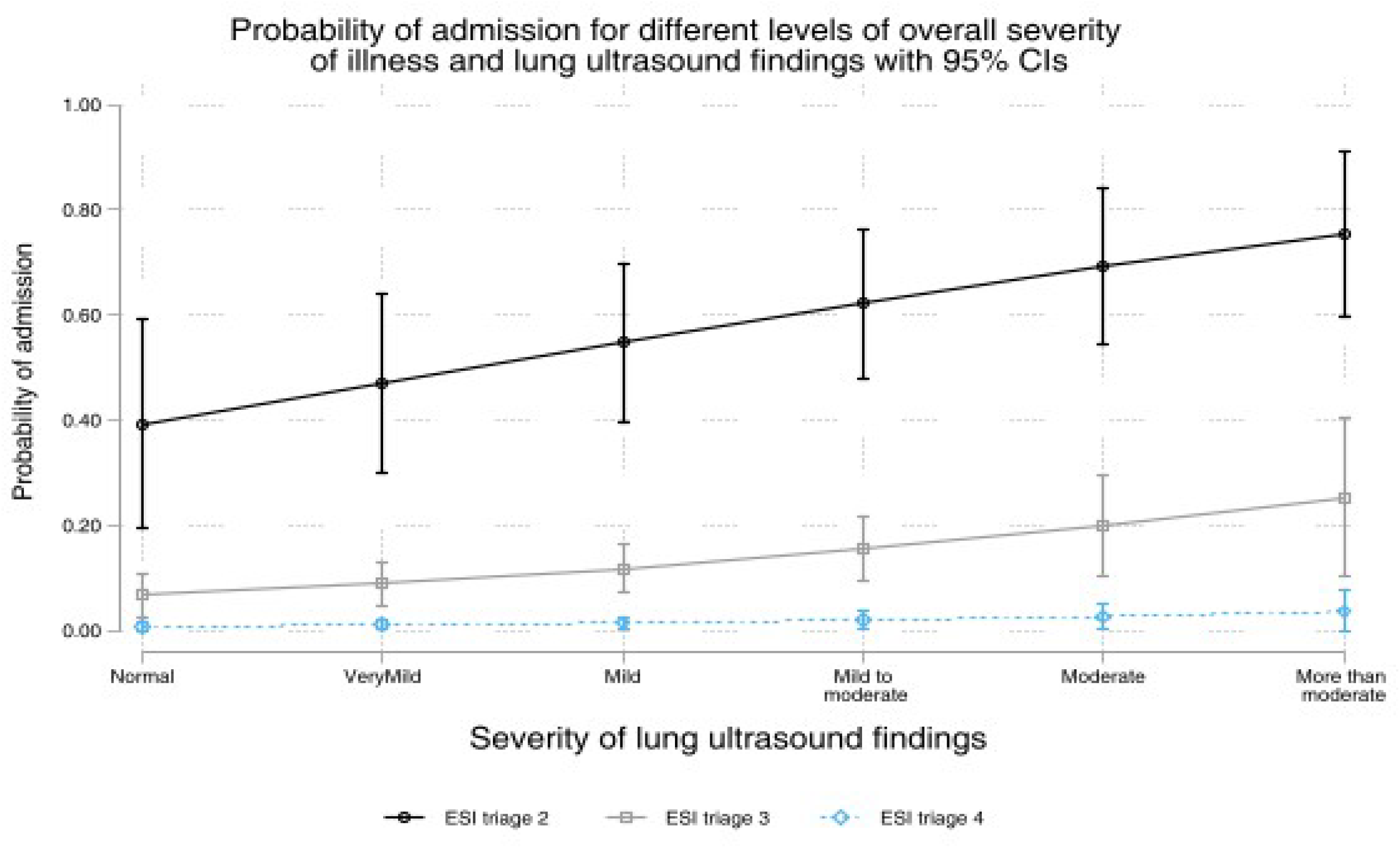
ESI, emergency severity index. The predicted probability of admission is calculated from the logistic regression model estimating the relationship between severity of lung ultrasound findings and hospital admission at different triage categories. The whiskers represent 95% confidence intervals.

### Blood tests

Eighty-four children who had lung ultrasounds performed had at least one laboratory blood test performed. The most commonly performed blood tests were the complete blood count (n=84) and C-reactive protein (n=70). The least commonly performed were the erythrocyte sedimentation rate (n=20) and pro-BNP (n=24). The combined categories of moderate to severe, and severe disease, comprised only 19 (4%) of the lung ultrasounds performed and it was only at this severity that laboratory abnormalities became apparent. The results of the blood tests for each category of lung ultrasound severity are in **Table 3**.

**Table 3.** Laboratory values observed by the severity of lung ultrasound, Median (Interquartile range). LDH lactate dehydrogenase, AST aspartate aminotransferase, ALT alanine transaminase, ESR erythrocyte sedimentation rate, CRP c-reactive protein, WBC white blood cell count, BNP b-natriuretic peptide, OR odds ratio, NC not calculable, LUS lung ultrasound, iu/l international units per litre, K/uL thousands per cubic millimeter, NAAT Nucleic acid amplification test.

| Factor | Normal | Very Mild | Mild | Mild to moderate | Moderate | More than moderate | Test of trend<br>Jonckheere–Terpstra |
| --- | --- | --- | --- | --- | --- | --- | --- |
| N | 200 | 68 | 68 | 30 | 42 | 19 | p-value |
| **CRP**<br>µg/mL | 13.6<br>(5.15, 29.75) | 19.65<br>(4.5, 51.65) | 7.5<br>(6.1, 7.8) | 13.55<br>(6.4, 36.7) | 6.8<br>(3, 12.2) | 22.1<br>(7.7, 28.9) | 0.6786 |
| **ESR**<br>mm/hour | 49<br>(42, 72.5) | 13.5<br>(11, 19) | 33<br>(14, 62) | 23.5<br>(11, 51.5) | 9<br>(4, 30) | 52<br>(42, 62) | 0.3096 |
| **Procalcitonin**<br>ng/mL | 0.06<br>(.06, .07) | 0.08<br>(.05, .19) | 0.055<br>(.02, .11) | 0.08<br>(.04, 2.06) | 0.135<br>(.04, .27) | 0.14<br>(.11, .23) | 0.1190 |
| **Ferritin**<br>ng/mL | 141<br>(40, 214) | 91<br>(73, 126) | 165<br>(83, 192) | 125<br>(77, 164) | 67<br>(28, 545) | 571.5<br>(122.5, 1050) | 0.1067 |
| **LDH**<br>iu/L | 305.5<br>(252.5, 314.5) | 238.5<br>(203, 289.5) | 228<br>(203, 306) | 246.5<br>(209, 268.5) | 306<br>(245, 357) | 441<br>(311, 549) | 0.0548 |
| **AST**<br>iu/L | 26<br>(17, 32.5) | 25<br>(16, 46) | 18<br>(16, 35) | 23<br>(16, 30) | 21<br>(19, 56) | 56<br>(42, 65) | 0.0108 |
| **ALT**<br>iu/L | 26<br>(16.5, 31) | 43<br>(18, 55) | 17<br>(8, 25) | 21<br>(15, 57) | 32<br>(15.5, 54) | 64<br>(41, 71) | 0.0074 |
| **Albumin**<br>mg/ml | 3.75<br>(3.6, 4.1) | 3.95<br>(3.6, 4) | 3.8<br>(3, 4.1) | 4.0<br>(3.05, 4.45) | 3.7<br>(3.4, 3.9) | 3.6<br>(3.2, 3.8) | 0.1416 |
| **Hematocrit**<br>% | 38 (36.8, 39.6) | 38.2 (37, 45) | 37.55 (34.25, 40.05) | 39.9<br>(38.1, 44) | 37.2<br>(35.15, 39.9) | 41<br>(37.4, 45.6) | 0.7889 |
| **WBC**<br>K/uL | 9.7<br>(6.9, 13.8) | 8.5<br>(7.4, 11.2) | 9.05<br>(5.55, 14.1) | 10.4<br>(6.6, 15.6) | 9.15<br>(4.95, 11.6) | 11.1<br>(6.3, 11.4) | 0.8935 |
| **Lymphocyte** | 2.6 | 1.4 | 0.9 | 4.7 | 1.9 | 1.6 | 0.9496 |
| K/uL | (2.6, 2.6) | (1.3, 1.5) | (0.9,0 .9) | (2.4, 6.9) | (1.5, 2.6) | (1.3, 3.75) |  |
| **Neutrophil**<br>K/uL | 4.4<br>(4.4, 4.4) | 2.95<br>(2.2, 3.7) | 10.8<br>(10.8, 10.8) | 3<br>(2.5, 3.5) | 4.4<br>(2.7, 21.6) | 6.2<br>(4.35, 7.2) | 0.3149 |
| **Pro-BNP**<br>pg/ml | 170<br>(40, 2918) | 346.5<br>(142, 551) | 1400<br>(1400, 1400) | 78<br>(58, 354) | 232<br>(64, 1276) | 2394<br>(200, 9319) | 0.3183 |
| **Creatinine**<br>mg/dL | 0.71<br>(0.38, 0.88) | 0.52<br>(0.48, 0.58) | 0.77<br>(0.62, 0.86) | 0.68<br>(0.45, 0.83) | 0.34<br>(0.26, 0.81) | 0.62<br>(0.39,0 .96) | 0.5426 |
| **Troponin-I**<br>(elevated) | 3 (38%) | 1 (17%) | 0 (0%) | 1 (13%) | 4 (29%) | 0 (0%) | 0.2929 |
| **D-dimer**<br>(elevated) | 3 (43%) | 4 (80%) | 2 (40%) | 2 (29%) | 8 (62%) | 8 (100%) | 0.0497 |
| **SARS-Cov2**<br>**(NAAT present**<br>**nose)** | 45/187<br>(24%) | 17/63<br>(27%) | 22/67<br>(33%) | (3/28)<br>(11%) | 14/37<br>(38%) | 7/16<br>(44%) | 0.091 |

We found some association of trend between severity of lung ultrasound findings and AST, ALT, Ferritin, and elevated D-dimer. Using linear regression Ferritin, LDH, AST, ALT, and D-dimer were significantly associated with the severity of lung ultrasound findings. The raw data is shown graphically for selected lab values in **Figure 2**. The regression analysis is summarized in **Table 4**. The following blood tests failed the screening process to be included in the final analysis: ESR, CRP, Procalcitonin, Albumin, Hematocrit, WBC, Pro-BNP, Troponin-I. The full analyses are in the appendices.

**Figure 2.**
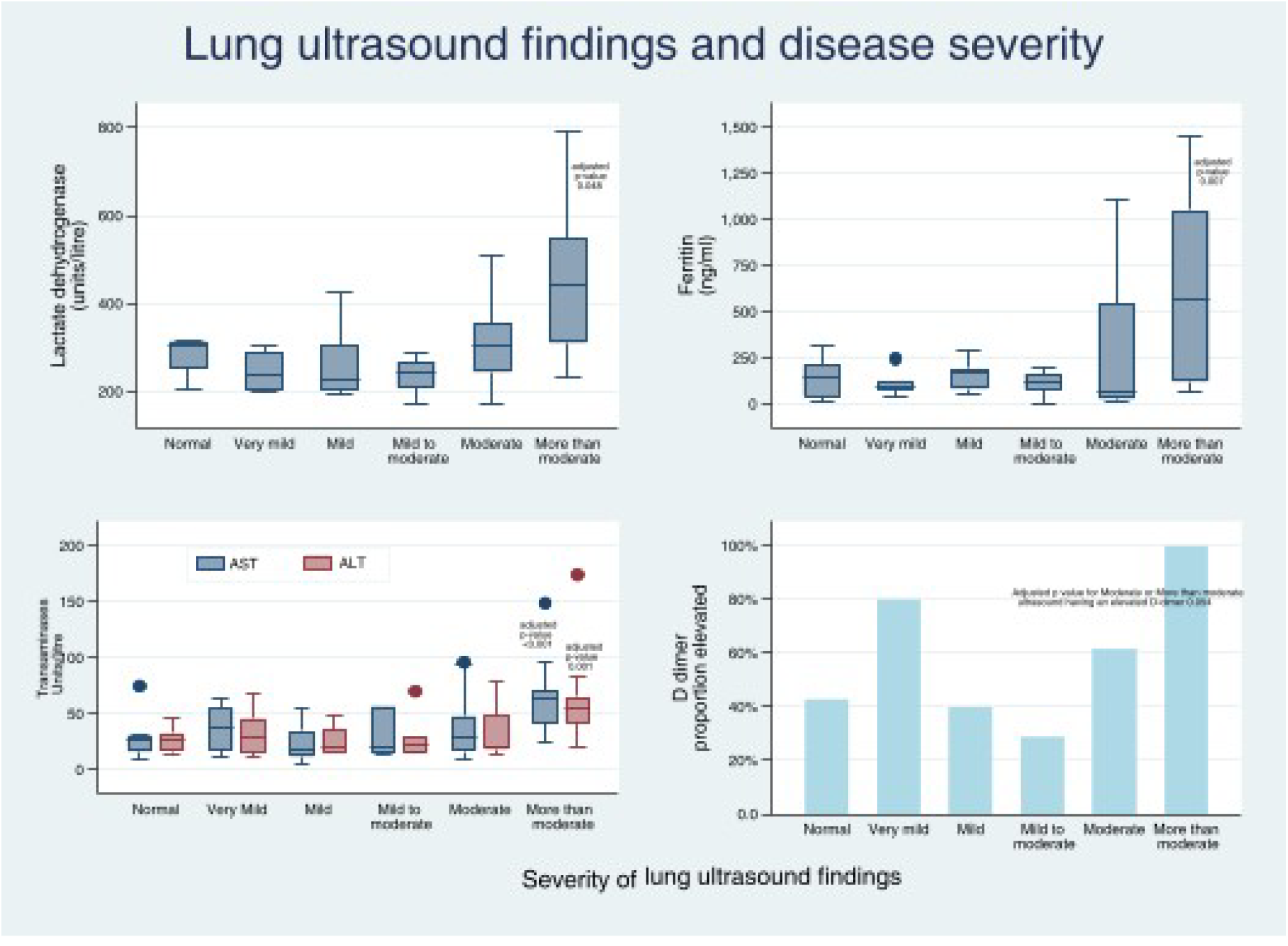
Barplots of lung ultrasound and disease severity. Except for the D-Dimer, important abnormalities in the markers shown are observed only for more than moderately severe lung disease. AST aspartate aminotransferase, ALT alanine transaminase.

**Table 4.**
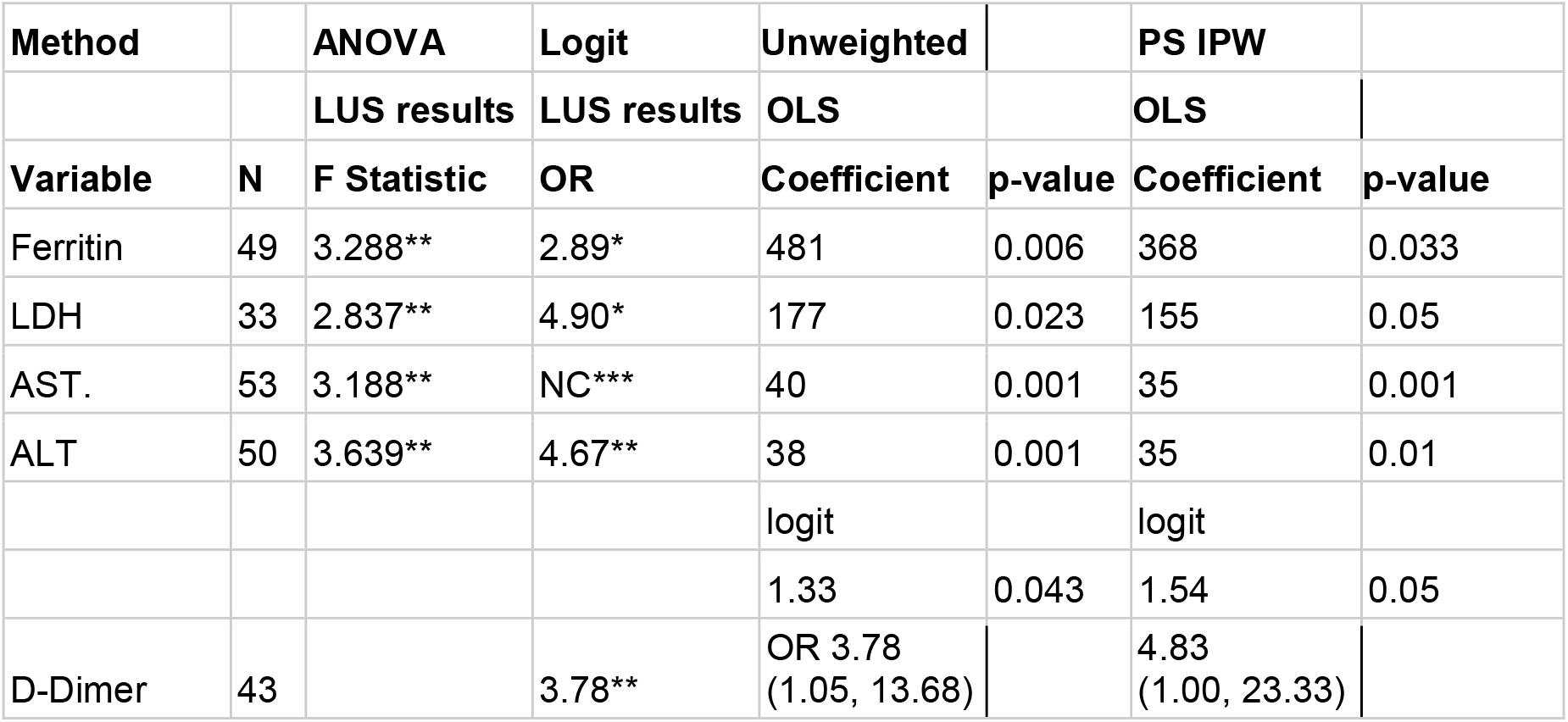
Models comparing the outcome to lung ultrasound severity. Laboratory tests had to pass a two-step process to be included in the final analysis. The first column shows the number of observations in each analysis. The second column shows the results of ANOVA relating ultrasound severity to mean laboratory values for each test. Where the p-value was <0.10 logistic regression relating lung ultrasound to the laboratory value being elevated was performed. These results are shown in the second column. If this result also had a p-value of <0.10 then that particular laboratory test was included in the final analysis. The fourth column shows ordinary least squares regression with laboratory value as the dependent variable and ultrasound severity as the independent variable. Ultrasound severity was treated as a categorical variable with normal as referent. Coefficients are shown for the ‘More than moderately severe’ ultrasound group. The fifth column shows the final result; the propensity score derived inverse probability-weighted ordinary least squares regression with laboratory value as the dependent variable and ultrasound severity as the independent variable Screening analysis *p<0.1, **p<0.05, ***p<0.001. Where ALT was reported only as <6 (n=3) it was not included in the analysis. Values for albumin ordered as a single add-on test by the admitting service were included in the analysis. D-dimer and troponin were analyzed only as elevated or normal. The full models are in the Appendix. OLS ordinary least squares, LDH lactate dehydrogenase, AST aspartate aminotransferase, ALT alanine transaminase, OR odds ratio, NC not calculable because all were abnormal in more severe ultrasound findings, LUS lung ultrasound, logit logistic regression, PS propensity score, IPW inverse probability-weighted.

In sensitivity analysis presented in **Table 5**, when we added age groups using schoolage children as the referent category we found relatively little effect although sample size limits the confidence of any assertions. The results suggest that LDH, AST, ALT, and ferritin tend to be slightly higher in younger children for a given severity of lung ultrasound.

**Table 5.** Sensitivity analysis showing the effect of age on laboratory values. Each column shows the results of an inverse probability-weighted logistic regression to see if age group affects the relationship between ultrasound findings and laboratory values. The beta coefficient reflects how the laboratory value would be modified for each age group for a given blood test. These results are not corrected for multiple comparisons. These models had poorer fit characteristics than the simpler regressions used in our main analysis. (*) indicates the laboratory value (primary independent variable) remained statistically significant following these adjustments performed in the sensitivity for age analysis. OR odds ratio.

| Age group | LDH |  | Ferritin |  | AST * |  | ALT* |  | D-dimer |  |
| --- | --- | --- | --- | --- | --- | --- | --- | --- | --- | --- |
| | $\beta$ | p | $\beta$ | p | $\beta$ | p | $\beta$ | p | OR | p |
| Neonate | 51 | 0.248 | 51 | 0.002 | 20 | 0.003 | 1 | 0.935 | 0.15 | 0.258 |
| Infant 1-12 months | 108 | 0.035 | 16 | 0.874 | 33 | 0.001 | 20 | 0.001 | 0.11 | 0.272 |
| Preschooler 1-5 years | 21 | 0.594 | -35 | 0.709 | 15 | 0.006 | 14 | 0.015 | 0.21 | 0.381 |
| School-age 6-12 years | Ref | - | Ref | - | Ref | - | Ref | - | Ref | - |
| Teenager 13-18 years | -47 | 0.153 | -17 | 0.773 | 6 | 0.416 | 8 | 0.283 | 0.08 | 0.083 |
| Transition 18-21 years | -8 | 0.859 | -17 | 0.825 | 6 | 0.475 | 6 | 0.461 | 0.25 | 0.395 |

## Discussion

We found that increasingly severe lung ultrasound abnormalities in children being evaluated for Covid in the ED were associated with increased admission rates. This helps validate lung ultrasound as a clinically useful tool. However, lung ultrasound findings were only part of the pathology leading to hospitalization in Covid-19 non-omicron waves, and lung injury alone as measured by lung ultrasound does not account for all admissions. This may appear self-evident, but anecdotal reports of Covid disposition being dichotomized to a specific SaO2 persist.

Some of the laboratory markers associated with poorer outcomes Ferritin, LDH, and elevated transaminases were particularly increased with ‘more than moderate’ severity lung disease suggesting a threshold effect on ultrasound. For D-dimer this was also true, but elevated D-dimers were also present in children with little or no disease on their lung ultrasounds. A normal lung ultrasound, in and of itself, is insufficient to decide to not obtain troponin, and Pro-BNP; ESR, CRP, and procalcitonin. Consequently, in children, while more than moderate pneumonitis on lung ultrasound should prompt sending blood tests, less severe or normal lung ultrasound findings on their own do not obviate the need to get blood tests.

Our finding that increasingly severe lung damage as demonstrated on bedside lung ultrasound is associated with an increased probability of hospitalization is unsurprising. It is consistent with CT findings in adult practice. However, our data also demonstrate that in children at least, even non-omicron strains of SARS-CoV-2 lead to hospitalization for reasons other than isolated lung injury. [21]

Our findings for laboratory abnormalities are broadly consistent with other studies. Similar associations to those we found have been described for lung ultrasound findings and ferritin levels [22] and lung CT and ferritin and D-dimer levels.[23] The consideration of Covid-19 as a new hyperferitinemic syndrome [6]has arisen from the observation that children who die frequently have respiratory and a hyperferritinemic manifestation of Covid. Post-mortem lung histology has shown evidence of elevated ferritin. Then again, autopsies are done on dead people, and our observation that lung damage and hyperferritinemia do not always go hand-in-hand is consistent with CT and pathological studies in adults.[24] Even in the presence of more severe lung pathology patients with a good prognosis have normal ferritin levels in contrast to those who do badly who tend to be hyperferritinemic.

In this mental model of Covid-19, although disease severity ranges from minimal symptoms to progressively more severe lung disease, the most severe form of Covid comprises severe lung disease plus the addition of a syndrome similar to macrophage activation syndrome (also termed secondary haemophagocytic lymphohistiocytosis). [11] This model, derived in adults, contemplates elevated ALT and AST in tandem with elevated ferritin,(typically >500 ng/ml[25]) and more severe lung injury.[11,26] This is what we observed here in our pediatric data also.

Omicron variants of SARS-CoV-2 have helped clarify the extrapulmonary pathogenicity of resulting Covid-19 in general. Accordingly, clinicians and health care planners must think beyond overly simplistic oxygen saturation thresholds as the only admission criterion as could occur if Covid-19 is characterized as requiring admission only if it causes hypoxia. Instead, management decisions must consider Covid-19 as having overlapping pulmonary, hematological, clotting, immunological, gastrointestinal, and cardiac components any one of which can lead to morbidity. Our work shows this is also true of non-omicron variants.

Myocarditis was identified early as a cause of death in adult patients and although death is rare in children myocarditis does occur. In our data, troponin elevations above the 99th centile were observed in 9/48 (19%) patients in whom they were drawn and bore no relation to lung ultrasound severity. Although not formally measured in our review, the decision to obtain troponin testing appears to have been guided by the appearance of irritability in preverbal and chest pain in verbal children as much as by lung ultrasound or clinical respiratory findings. Unlike adults, elevated troponin in children does not usually reflect ischemia or renal failure, and from the ED perspective has to be managed as reflecting myocarditis. ECGs are insufficiently sensitive to rule out myocarditis. A formal diagnosis of myocarditis requires a tissue diagnosis, immunohistological staining, and extensive serum antibody testing. [27] A practical working diagnosis of myocarditis for pediatric emergency medicine is one of irritability, diaphoresis with feeding, chest pain, palpitations, new heart murmur, hepatomegaly, edema, or cardiomegaly in combination with either a suggestive ECG, or CXR, or an elevated troponin. Using this framework, it is likely that all of these elevated troponins reflected myocarditis. Our findings are inconvenient for pediatric emergency practice; in an irritable infant with Covid-19, even in the absence of lung involvement troponin testing is still be indicated.

Our finding that D-dimer was universally elevated in those children with the most severe lung disease but could also be elevated in those without lung disease provides support for the concept of distinct processes of lung centric pulmonary intravascular coagulation [11] and peripheral sepsis-induced disseminated intravascular coagulation[28] that could occur separately or simultaneously.

In short, imaging the lungs informs the clinician about the lungs. A conceptual model of Covid-19 disease where a hyperferritinemic state with its associated laboratory abnormalities, morbidity and mortality complicate some, but not all, patients with more severe lung disease is emerging. This conceptual model dovetails with our findings. Lung ultrasound does not inform the clinician about the presence of myocarditis or peripheral inflammation.

Operationalizing our results requires the incorporation of information beyond this research. First, if a child has more than moderately severe pneumonitis on lung ultrasound ferritin and other markers of severe disease should be obtained. For lesser degrees of lung ultrasound abnormalities, the decision to obtain laboratory testing will rest on other clinical factors such as chest pain or irritability and in these cases, testing should then focus on troponin and inflammatory markers.

D-dimer results reflect both pulmonary and peripheral clotting and the decision to test rests on the philosophy of treating physicians regarding the management of apparent pulmonary and disseminated hypercoagulable states in children.

## Limitations

Our sample size is small and reflects the experiences of a single center. The small sample size risks sparse data problems for variables with binary outcomes such as D-dimer which, because of a change in assays during the study period could only be treated as elevated or normal. We treated each visit rather than the patient as the unit of analysis. We had a small number of ultrasound operators. This helps ensure consistency of the scans performed and their interpretation but potentially at the price of generalizability. The sample is also non-random; the high proportion testing positive on nasal swabbing for SARS-CoV-2 suggests lung ultrasound was used rather selectively. Given the airborne transmission of SARS-CoV-2, many patients with pneumonitis on lung ultrasound likely had SARS-CoV-2 infection, particularly given the number of patients with no alternative viruses detected. However, definitively proving or disproving lower respiratory tract SARS-CoV-2 infection is not possible in most clinical settings. From a clinical practice perspective, the combination of a negative nasal swab and lung imaging is as close as one is likely to get to objectively declare a patient Covid free.

The inherent difficulty in case definition is less of a limitation in emergency medicine research than other areas of research because pediatric emergency physicians must generally make their management decisions (including whether to obtain blood tests and often disposition) before PCR test results are available. The results of the lung ultrasound and history and physical exam findings are, by contrast, immediately available.

We did not use an ultrasound scoring system. Such systems are inherently attractive when developing prospective research. However, they are often time-consuming to use and this can lead to patients being inappropriately excluded from studies. Even if this pitfall can be avoided, the scores generated by scoring schemas can create an illusion of precision. There was some evidence of a similar effect even in the absence of a scoring system; the ultrasonographers in this study interpreted far more categories of severity of lung ultrasound than the subsequent analysis which correlated these with laboratory abnormalities supported.

Our sample includes a broad age range of patients. We addressed this with a sensitivty analysis to answer the question did age group matter? This method allows us to avoid the problem of subdividing our sample into many smaller groups each of which contains insufficient data to inform and may prove useful in pediatric research in general. Our sensitivity analyses suggest that LDH, AST, ALT, and ferritin tend to be slightly higher in infants and younger children compared to older children for a given severity of lung ultrasound; although the magnitude was small. These findings are consistent with the findings of a pooled analysis of studies of laboratory abnormalities and disease severity in children with Covid-19. [8] Our findings with respect to leukocyte findings in children add only to the diverse and inconsistent findings described by many others.

Covid 19 in the pediatric population is generally milder than in the adult ED population. Consequently, research in children may allow researchers to tease out the different cardiac, hematological, clotting hyperferritinemic, and lung components of Covid-19 disease that are otherwise difficult to separate in adults because of intervening morbidity and mortality. Where some will see this as an opportunity for a detail-oriented practice precisely delineating the manifestations of Covid-19 to prognosticate for each child, others will favor a minimalist approach reserving lung ultrasound and other time-consuming investigations only for those children who are in severe overt distress at the time of presentation.

In summary, more severe lung ultrasound abnormalities were associated with more hospitalization, and more than moderate severity of lung ultrasound abnormalities were associated with increased hyperferritinemia, elevated LDH, elevated transaminases, and elevated D-dimer. A normal lung ultrasound does not obviate the need to obtain troponin levels or D-dimer levels. Lung pathology alone did not fully account for the hospitalization of children with Covid-19 even in pre-omicron waves.

## Data Availability

Data is available from a data steward at Sutter Institute for Medical Research subject to their rules.

## Appendices

Appendix Full respiratory virus panel results

Appendix Full regression models (code)

Appendix Sample of US reports and their classification

## Notes

### Competing Interest Statement

The authors have declared no competing interest.

### Funding Statement

The Pediatric Emergency Medicine Research Foundation and Sutter Medical Center Foundation Sacramento

### Author Declarations

IRB of Sutter Health gave ethical approval for this work.

