## Appendices for "More severe pneumonitis in children predicts the need for admission and elevation of some but not all markers of severe Covid-19"

Appendix 1. SQL Code

<Pending release from data steward>

Appendix 2. Viral testing

| Covid testing | N | % |
|---|---|---|
| Negative | 290 | 68 |
| Positive | 108 | 25 |
| Not tested | 29 | 7 |
| Total | 427 | 100 |

| Viral Panel | Freq. | Percent |
|---|---|---|
| CORONAVIRUS HKU1 | 2 | 0.5 |
| CORONAVIRUS NL63 | 5 | 1.2 |
| CORONAVIRUS OC43 | 3 | 0.7 |
| CORONAVIRUS 229E | 2 | 0.5 |
| RHINOVIRUS/ENTEROVIRUS | 73 | 17.1 |
| INFLUENZA A | 4 | 0.9 |
| INFLUENZA B | 1 | 0.2 |
| PARAINFLUENZA 1 VIRUS | 1 | 0.2 |
| PARAINFLUENZA 3 VIRUS | 6 | 1.4 |
| CHLAMYDOPHILA PNEUMONIAE | 1 | 0.2 |
| SARS-CoV-2 (including dual diagnosis) | 108 | 25.3 |
| No Virus found | 221 | 51.8 |
| Total | 427 | 100.0 |

Appendix 3. Regression Models

| | | | | | | | | | |
|---|---|---|---|---|---|---|---|---|---|
| Variable N | | ANOVA | Logit | **Unweighted OLS** | | Parsimonious | | Detailed | |
| | | LUS results | LUS results | | | **Inverse probability OLS** | | **Inverse probability OLS** | |
| | | F Statistic | OR | **Coefficient** | **p-value** | **Coefficient** | **p-value** | **Coefficient** | **p-value** |
| Ferritin | 49 | 3.288** | 2.89* | 481 | 0.006 | 478 | 0.007 | 368 | 0.033 |
| LDH | 33 | 2.837** | 4.90* | 177 | 0.023 | 167 | 0.047 | 155 | 0.05 |
| AST. | 53 | 3.188** | NC*** | 40 | 0.001 | 38 | 0.001 | 35 | 0.001 |
| ALT | 50 | 3.639** | 4.67** | 38 | 0.001 | 43 | 0.004 | 35 | 0.01 |
| | | | | logit | | logit | | logit | |
| | | | | 1.33 | 0.043 | 1.54 | 0.05 | | |
| D-Dimer | 43 | | 3.78** | OR 3.78 (1.05, 13.68) | | 4.83 (1.00, 23.33) | | | |

This summarizes the results using competing modelling strategies for the propensity score models. Balance graphs show the effects on balance. Full models are shown in the code following the graphs. LUS lung ultrasound

Variables that failed screening steps to be included in the analysis:

| Variable | N | ANOVA<br>F | Logit<br>Odds ratio |
|---|---|---|---|
| ESR | 27 | 2.647* | 0.64 |
| CRP | 61 | 0.299 | |
| Procalcitonin | 55 | 0.473 | |
| Albumin | 58 | 0.644 | |
| Hematocrit | 58 | 0.585 | |
| WBC | 88 | 0.056 | |
| Pro-BNP | 21 | 0.855 | |
| Troponin-I | 48 | | 0.80 |

*P<0.05

Effect of propensity scores on balance

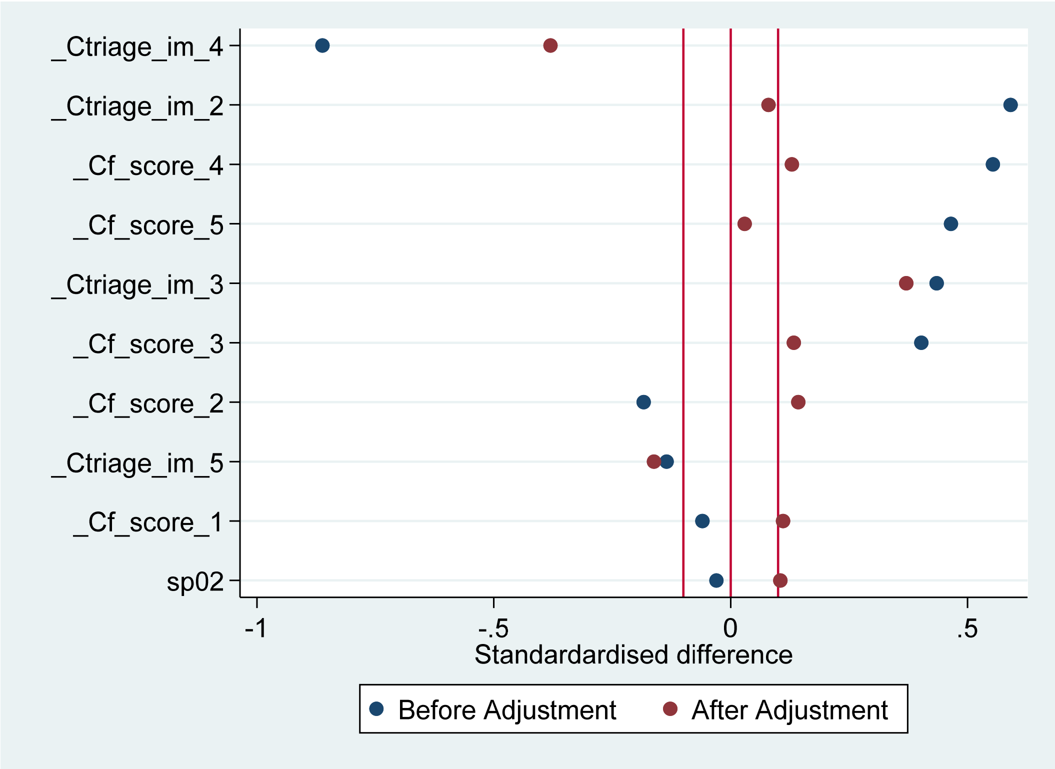

Effect of propensity score weighting on balance – Ferritin parsimonious

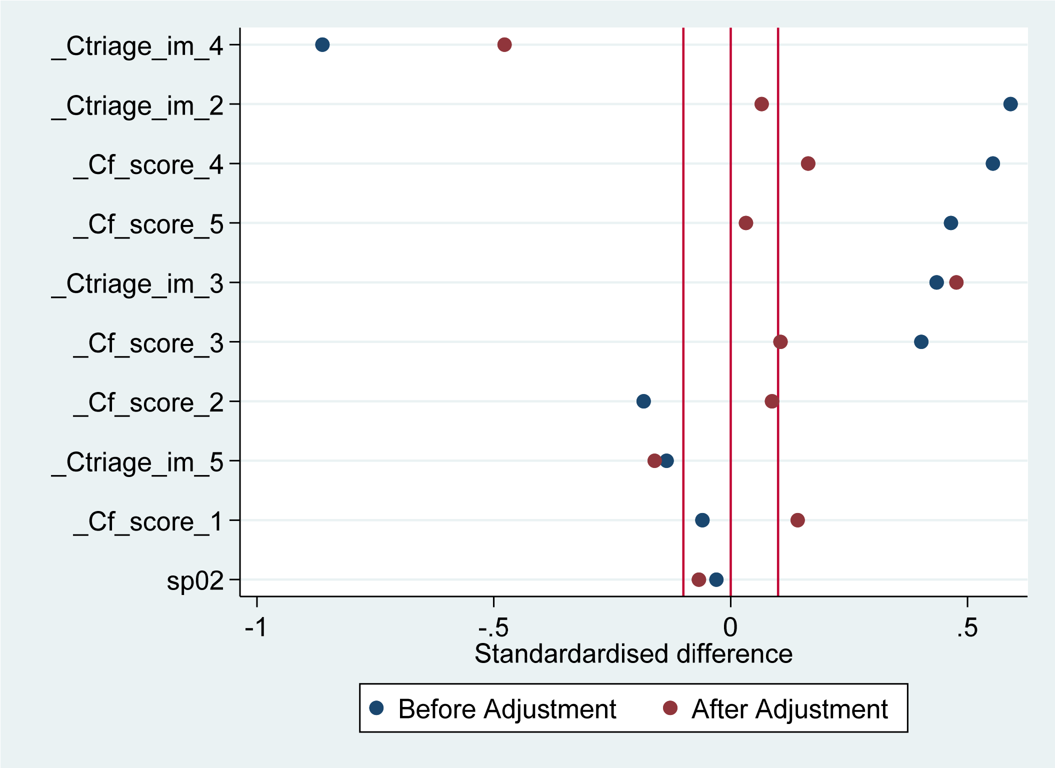

Effect of propensity score weighting on balance – Ferritin detailed

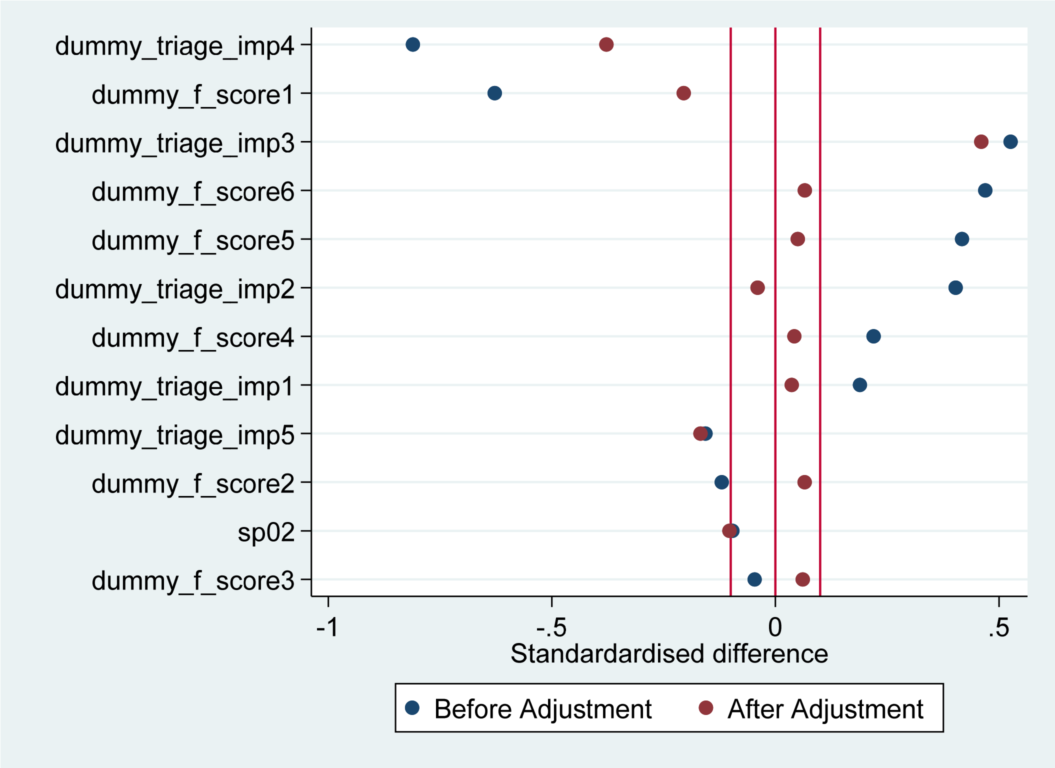

Effect of propensity score weighting on balance – AST parsimonious

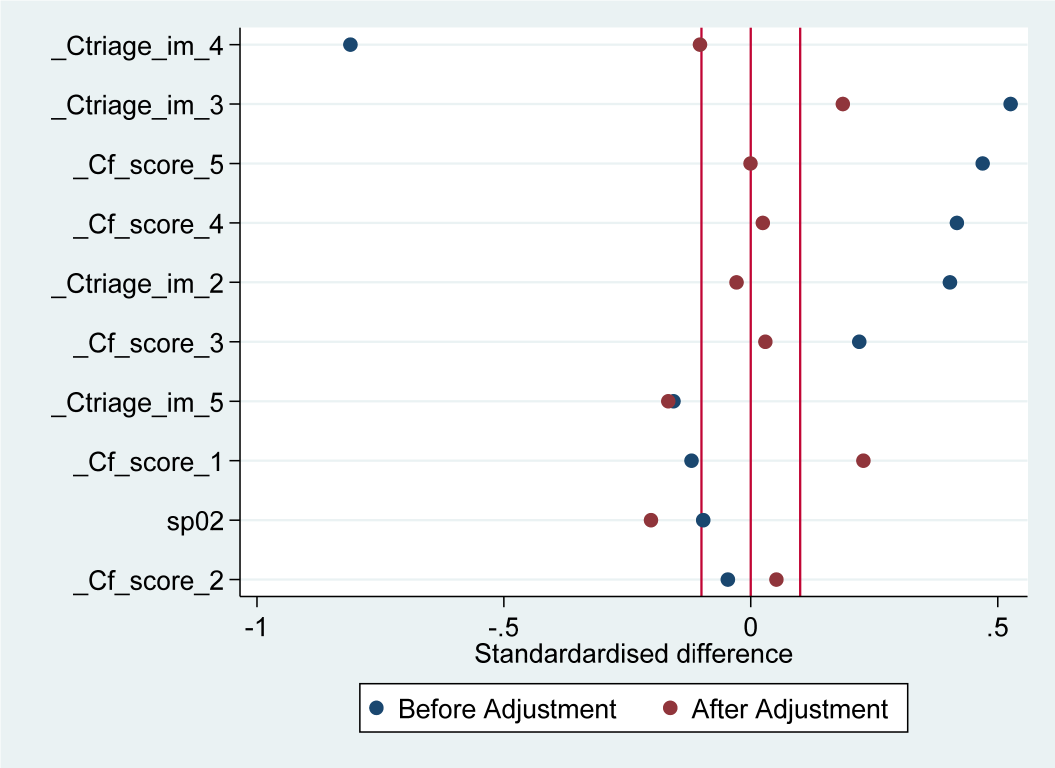

Effect of propensity score weighting on balance – AST detailed

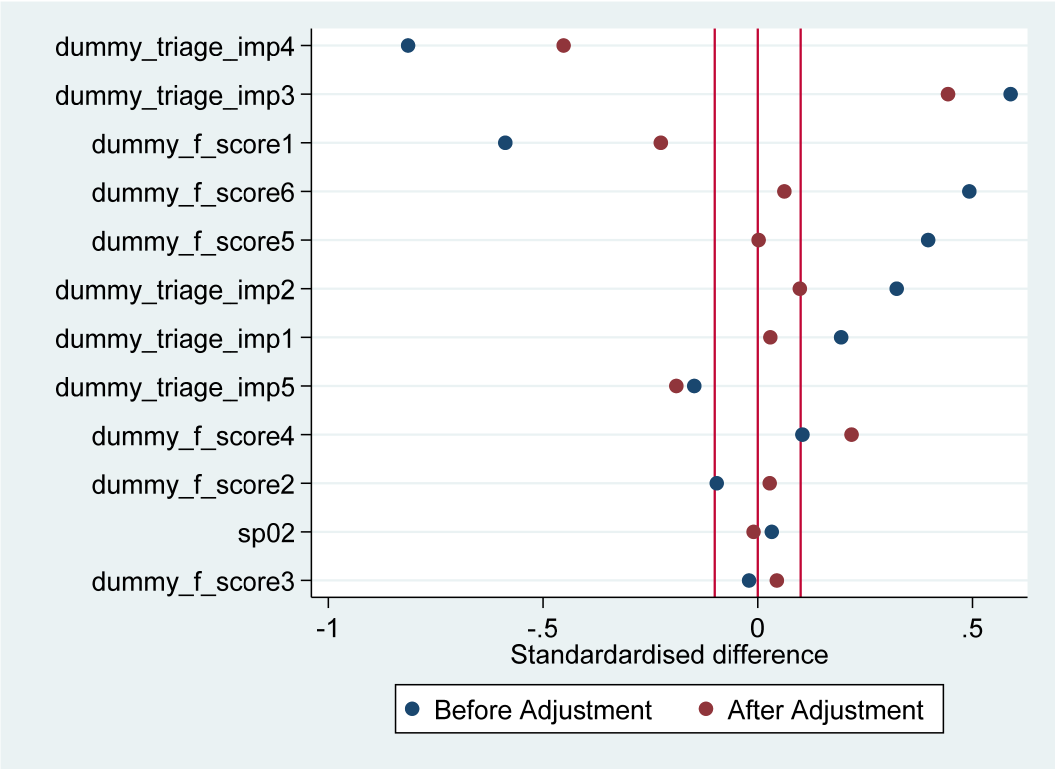

Effect of propensity score weighting on balance –ALT Parsimonious

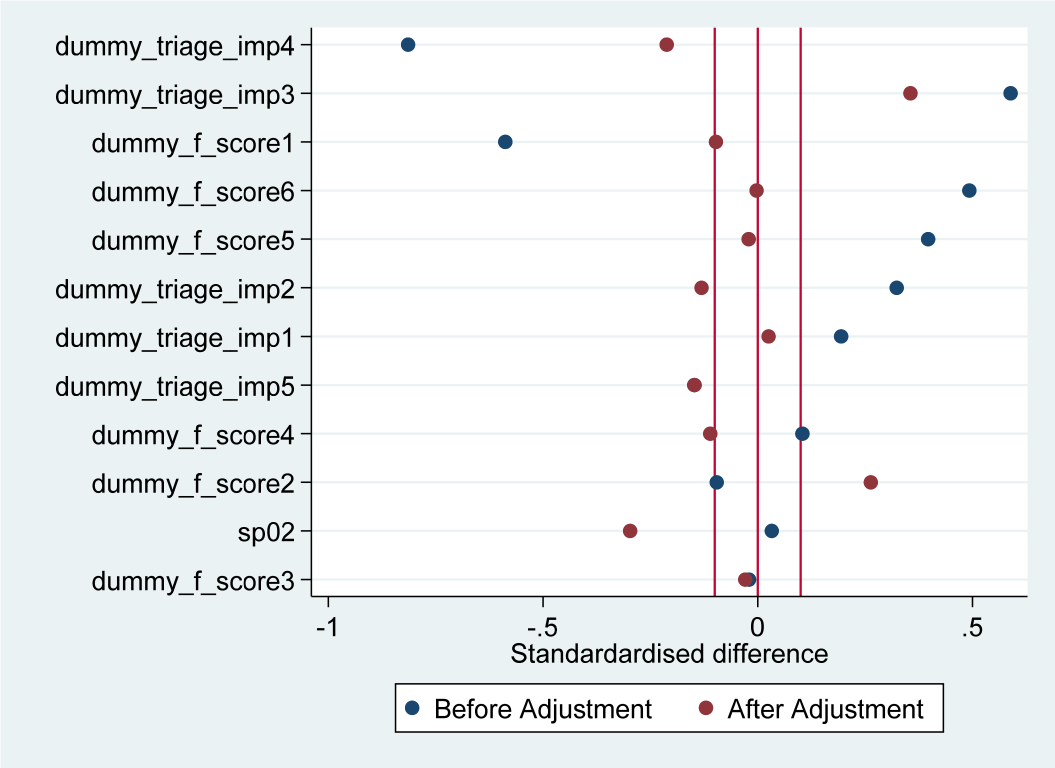

Effect of propensity score weighting on balance –ALT detailed

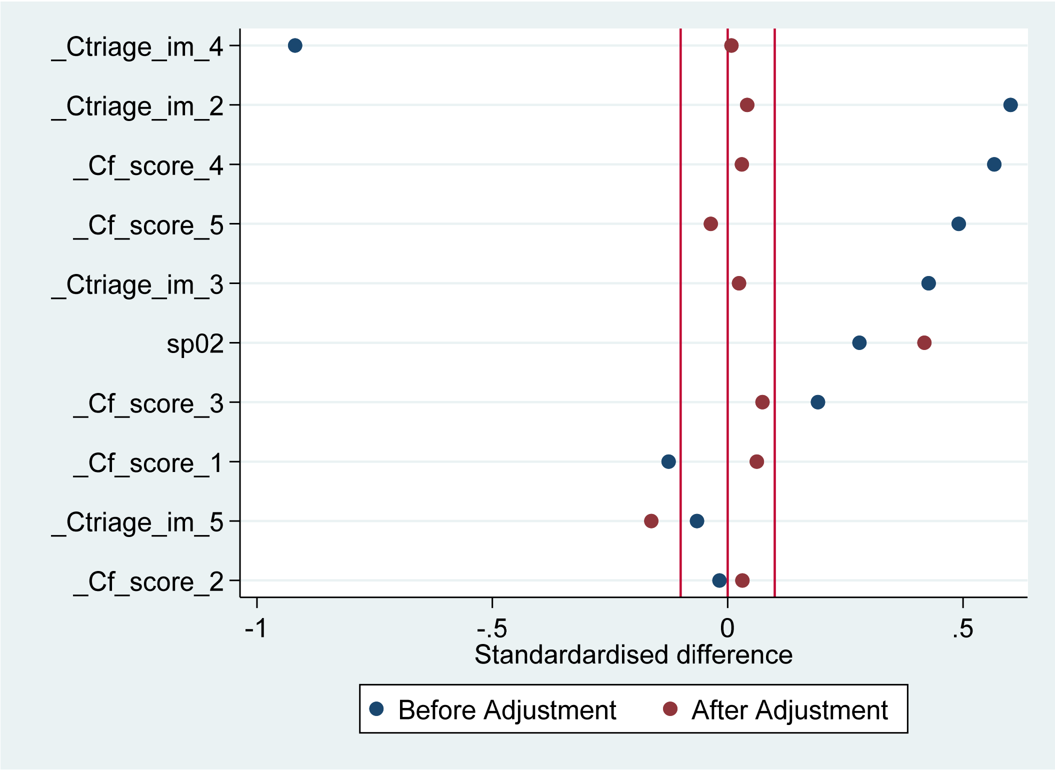

Effect of propensity score weighting on balance –LDH – Parsimonious model

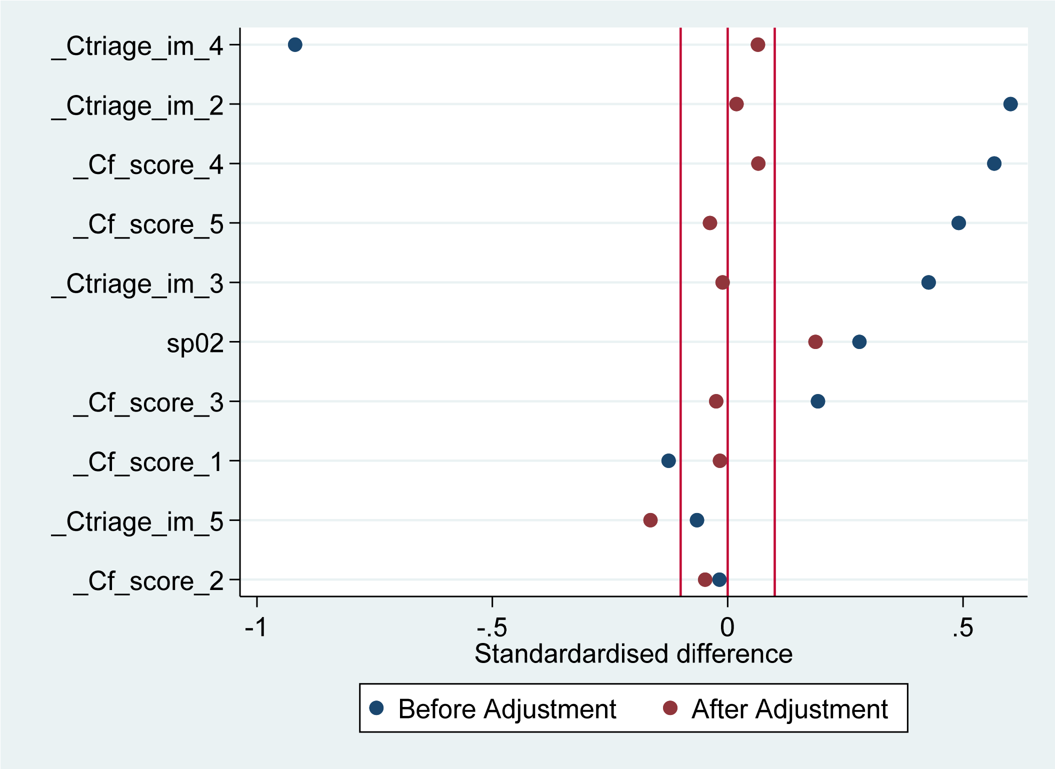

Effect of propensity score weighting on balance –LDH – Detailed model

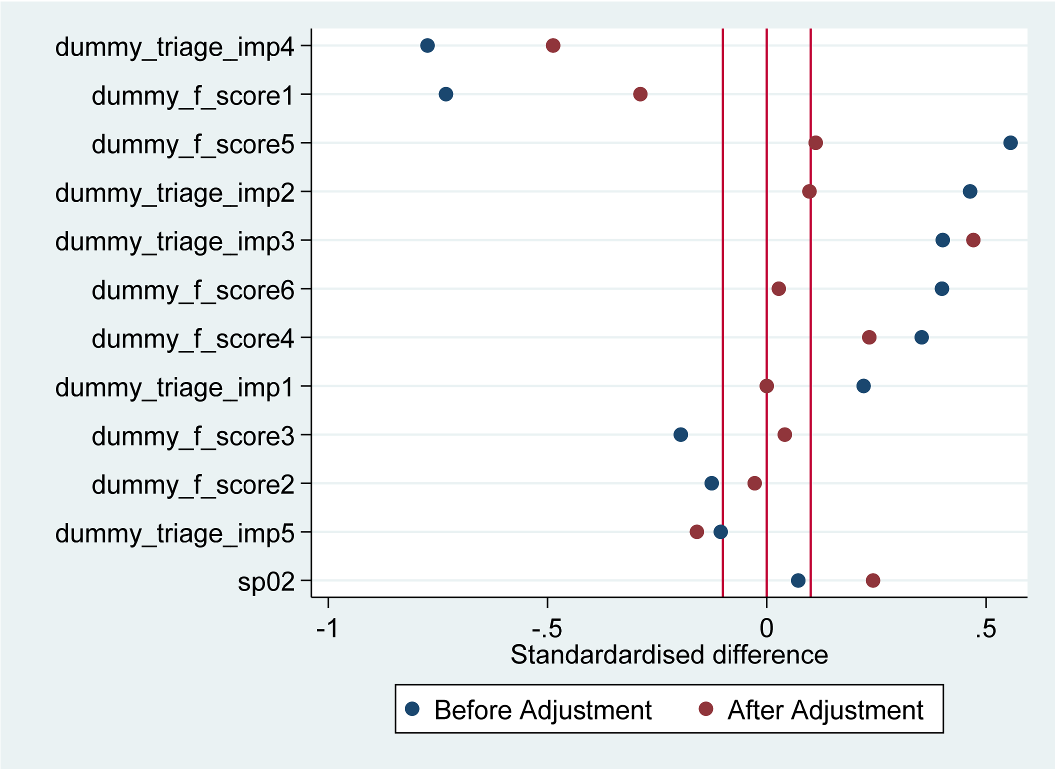

Effect of propensity score weighting on balance D-Dimer parsimonious

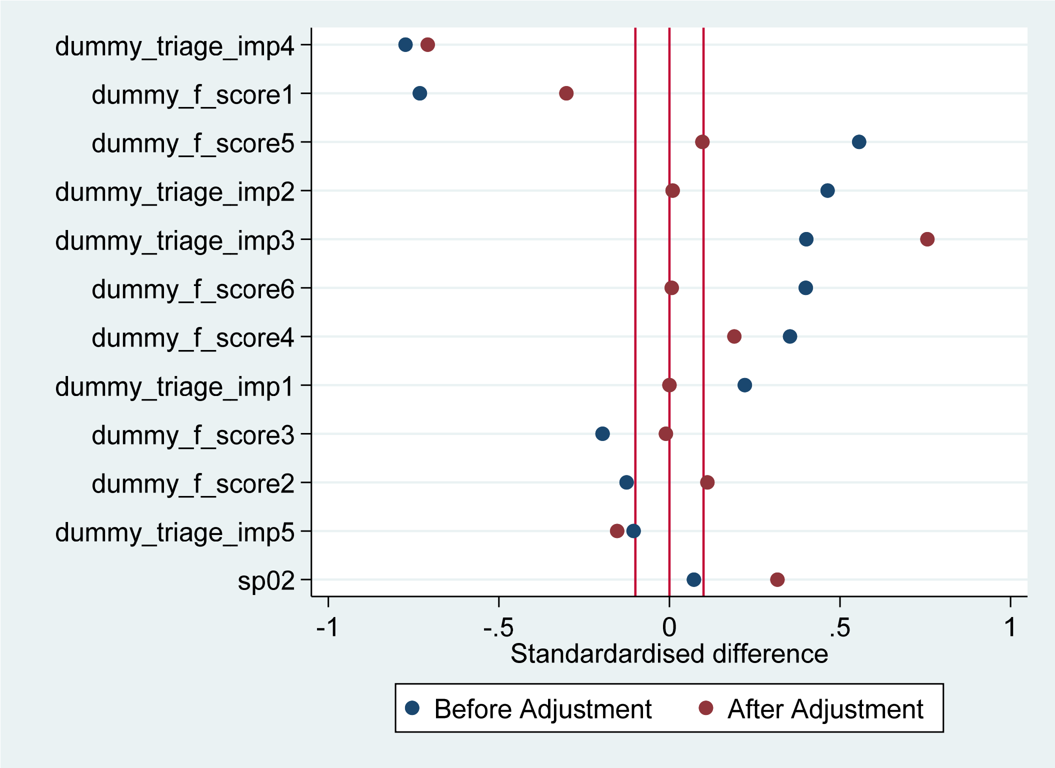

Effect of propensity score weighting on balance D Dimer detail [unlike other parsimonious is as good/better]

[uses T16.dta]

Propensity score part

```
**# Bookmark #3
//ferritn parsimonious
 foreach var in ferritin_measure {
            cap drop  iptwt_`var'
             logit `var' i.f_score i.triage_imp o.admit
             estat gof
             cap drop propensity
              predict propensity ,pr

              propwt ferritin_measure propensity, ipt gen(wt_`var')

              reg l_ferritin i.f_score

              reg l_ferritin i.f_score [pweight =iptwt_`var']
 }
    xi: pbalchk  ferritin_measure   i.f_score  i.triage_imp sp02  ,wt(iptwt_ferritin_measure  ) graph

//ferritin detailed

cap drop reg_l_fer_norm
cap drop reg_l_fer_propwt
 foreach var in ferritin_measure {
            cap drop  iptwt_`var'

             logit `var' i.f_score i.triage_imp   sp02 age_y
             estat gof
             cap drop propensity
```

```
        predict propensity ,pr

        propwt ferritin_measure propensity, ipt gen(wt_`var')

        reg l_ferritin i.f_score
          predict reg_l_fer_norm ,xb

        reg l_ferritin i.f_score [pweight =iptwt_`var']
          predict reg_l_fer_propwt ,xb
}
 tw (lowess reg_l_fer_norm f_score )(sc l_ferritin  f_score ) (lowess reg_l_fer_propwt  f_score ) (lowess l_ferritin
f_score )

  xi: pbalchk  ferritin_measure   i.f_score  i.triage_imp sp02  ,wt(iptwt_ferritin_measure  ) graph

 //AST Parsimonious
  cap drop ast_done
gen ast_done =1 if l_lft_ast <.
replace ast_done =0 if l_lft_ast ==.

        foreach var in ast_done {
       cap drop  iptwt_`var'
        logit `var' i.f_score##c.triage_imp o.admit
        estat gof
        cap drop propensity
        predict propensity ,pr

        propwt ast_done  propensity, ipt gen(wt_`var')

        reg l_lft_ast i.f_score
```

```
        reg l_lft_ast  i.f_score [pweight =iptwt_`var']
}
  xi: pbalchk  ast_done   dummy_f_score* dummy_triage_imp*    sp02  ,wt(iptwt_ast_done  ) graph
```

//AST detailed

```
cap drop ast_done
gen ast_done =1 if l_lft_ast <.
replace ast_done =0 if l_lft_ast ==.

        foreach var in ast_done {
       cap drop  iptwt_`var'
        logit `var' i.f_score c.f_score i.triage_imp sp02 c.age_y##c.triage_imp
        estat gof
        cap drop propensity
        predict propensity ,pr

        propwt ast_done  propensity, ipt gen(wt_`var')

        reg l_lft_ast i.f_score

        reg l_lft_ast  i.f_score [pweight =iptwt_`var']
}

 xi: pbalchk  ast_done   i.f_score  i.triage_imp sp02  ,wt(iptwt_ast_done  ) graph
```

//ALT Parsiominious

```
 cap drop alt_done
gen alt_done =1 if l_lft_alt <.
replace alt_done =0 if l_lft_alt ==.

               foreach var in alt_done {
             cap drop  iptwt_`var'
              logit `var' i.f_score##c.triage_imp i.admit
              estat gof
              cap drop propensity
               predict propensity ,pr

               propwt alt_done propensity, ipt gen(wt_`var')

               reg l_lft_alt i.f_score

               reg l_lft_alt  i.f_score [pweight =iptwt_`var']
}

xi: pbalchk  alt_done   dummy_f_score* dummy_triage_imp*    sp02  ,wt(iptwt_alt_done  ) graph

//ALT Detailed

 cap drop alt_doneA
gen alt_doneA =1 if l_lft_alt <.
replace alt_doneA =0 if l_lft_alt ==.

               foreach var in alt_doneA {
             cap drop  iptwt_`var'
              logit `var' i.f_score##c.triage_imp   sp02 c.age_y##c.triage_imp
              estat gof
              cap drop propensity
```

```
            predict propensity ,pr

            propwt alt_doneA propensity, ipt gen(wt_`var')

            reg l_lft_alt i.f_score

            reg l_lft_alt  i.f_score [pweight =iptwt_`var']
}
xi: pbalchk  alt_doneA   dummy_f_score* dummy_triage_imp*    sp02  ,wt(iptwt_alt_doneA  ) graph

           //parsimonious
                  foreach var in ldh_done {
           cap drop  iptwt_`var'
           logit `var'   i.f_score i.triage_imp

           estat gof
           cap drop propensity
            predict propensity ,pr

            propwt ldh_done propensity, ipt gen(wt_`var')

            reg l_ldh i.f_score

            reg l_ldh  i.f_score [pweight =iptwt_`var']
}
xi: pbalchk  ldh_done   i.f_score  i.triage_imp sp02  ,wt(iptwt_ldh_done  ) graph

  //detailed
```

```
 foreach var in ldh_done {
cap drop  iptwt_`var'
 logit `var'   i.f_score i.triage_imp    sp02 c.age_y c.triage_imp

 estat gof
 cap drop propensity
  predict propensity ,pr

  propwt ldh_done propensity, ipt gen(wt_`var')

  reg l_ldh i.f_score

  reg l_ldh  i.f_score [pweight =iptwt_`var']
}
xi: pbalchk  ldh_done   i.f_score  i.triage_imp sp02  ,wt(iptwt_ldh_done  ) graph

 foreach var in dimer_measured {
cap drop iptwt_`var'
 logit `var'   i.f_score i.triage_imp pts_that_hourP21

  estat gof
 cap drop propensity
  predict propensity ,pr

  propwt dimer_measured propensity, ipt gen(wt_`var')

 logit elev_dimer f456, or

 logit elev_dimer f456 i.age_group ,or
```

```
logit elev_dimer f456 [pw =iptwt_`var'] ,or

logit elev_dimer f456 i.age_group [pw =iptwt_`var']  ,or
}
xi: pbalchk  dimer_measured   dummy_f_score*  dummy_triage_imp*  sp02  ,wt(iptwt_dimer_measured  ) graph
```

// The issue here is that F56 has perfect predcition for 8 values. P values are very unstable and can get well below 0.05 using exlogisitc and manually generated weighted outcomes. But, F456 avoids perfect prediction and seems to proved estimates that appeart more reasonable.

```
//detailed
  foreach var in dimer_measured {
 cap drop iptwt_`var'
    logit `var'   i.f_score i.triage_imp##c.sp02 c.age_y c.triage_imp pts_that_hourP14

     estat gof
    cap drop propensity
     predict propensity ,pr

     propwt dimer_measured propensity, ipt gen(wt_`var')

    logit elev_dimer f456, or

    logit elev_dimer f456 i.age_group ,or

    logit   elev_dimer f456 [pw =iptwt_`var'] ,or

    logit elev_dimer f456 i.age_group [pw =iptwt_`var']  ,or
}

xi: pbalchk  dimer_measured   dummy_f_score*  dummy_triage_imp*  sp02  ,wt(iptwt_dimer_measured  ) graph
```

```
**## Table 3  Raw lab data for each category

// See spreadsheet for code

loc flagD "if flagD==0"

table1 `flagD' ,by(f_score) vars(l_crp conts\  l_esr  conts\l_procalcitonin conts\ l_ferritin  conts\ l_ldh conts\ l_lft_alt conts\ l_lft_ast conts\ l_alb  conts\ l_hematocrit  conts\ l_cbc_wbc conts\ l_abs_lymphocyte conts\ l_abs_neutrophil conts\  l_bnp conts\ l_creat conts\ elev_trop bine \elev_dimer bine)   saving(table_3b_flagD.xlsx ,replace)

//table_1mc allows total column

**# Figure 2 Graph (components first)

 gr box l_ldh ,over(f_score )

   gr play ldh_graph.grec

    graph save "Graph" "E:\2022_1_January\Covid stuff\ldh_graph.gph"

  gr box l_ferritin ,over(f_score )

   // manual clean up in editor

    graph save "Graph" "E:\2022_1_January\Covid stuff\ferritin_graph.gph"

         gr box l_lft_ast l_lft_alt ,over(f_score )

         //manual clean up

         graph save "Graph" "E:\2022_1_January\Covid stuff\AST_ALT_graph.gph", replace

         graph save "Graph" "E:\2022_1_January\Covid stuff\dimer_graph.gph"

     graph save "Graph" "E:\2022_1_January\Covid stuff\dimer_graph.gph", replace

         //////////
```

```
	//Create a marker for US experience by operator
	//

	///

**#   Used in seprate do file to pull ot the Sp02 then merged back in.
/*
cap drop t1
cap drop t2
cap drop t3
cap drop t4
cap drop t5
cap drop t6
cap drop t7

gen t1 =strpos(note_text, "SpO2")

gen t3 = substr(note_text ,t1 ,35)

gen t4 =(strpos(t3, "%")  -4)
gen t5 =substr(t3,t4,4)

gen t6 = .
replace t6 =100 if regexm(t5, "100")
replace t6 =99 if regexm(t5 ,"99")
replace t6 =98 if regexm(t5 ,"98")
replace t6 =97 if regexm(t5 ,"97")
replace t6 =96 if regexm(t5 ,"96")
replace t6 =95 if regexm(t5 ,"95")
replace t6 =94 if regexm(t5 ,"94")
replace t6 =93 if regexm(t5 ,"93")
replace t6 =92 if regexm(t5 ,"92")
```

```
replace t6 =91 if regexm(t5 ,"91")
replace t6 =90 if regexm(t5 ,"90")
replace t6 =89 if regexm(t5 ,"89")
replace t6 =88 if regexm(t5 ,"88")
replace t6 =87 if regexm(t5 ,"87")
replace t6 =86 if regexm(t5 ,"86")
replace t6 =85 if regexm(t5 ,"85")
replace t6 =84 if regexm(t5 ,"84")
replace t6 =83 if regexm(t5 ,"83")
replace t6 =82 if regexm(t5 ,"82")
replace t6 =81 if regexm(t5 ,"81")
replace t6 =80 if regexm(t5 ,"80")

replace t6 =.a if regexm(t5, "/=")
replace t6 =.b if regexm(t3, "Min:")
replace t6 =.b if regexm(t3, "Max")

        */

**# Sp02 Groupings
cap drop gr_sp02
cap lab drop o2

gen gr_sp02 =1 if sp02 >=97
replace gr_sp02 =2 if sp02 <97 & sp02>=94
replace gr_sp02 =3 if sp02 <94 & sp02>=92
replace gr_sp02 =4 if sp02 <92 & sp02>=90
replace gr_sp02=5 if sp02 <90
replace gr_sp02 =6 if sp02 <=85
```

```
lab define ox 1 "sp02 >=97" 2 "sp02 94%-96%"  3 "Sp02 92%-94%" 4 "Sp02 91%-90%" 5 "Sp02 <90%"  6 "Sp02 <85%"

lab val gr_sp02 o2

loc flagD "if flagD==0"

// Paragraph 3 Results  Admission
nptrend admit `flagD'  ,gr(f_score ) carmitage
nptrend admit `flagD' ,gr(triage_imp ) carmitage
nptrend admit `flagD' ,gr(gr_sp02  ) carmitage

logit admit  f_score  triage_imp `flagD' ,or
margins ,at(f_score =(0/5))
marginsplot

//

//

// Table 3  Test of trend column
//
foreach var in l_crp l_abs_lymphocyte l_abs_neutrophil l_al l_bicarb l_bnp l_cbc_wbc l_creat l_dimer_new l_dimer_old l_esr l_ferritin l_hematocrit l_ldh l_lft_ast l_lft_alt l_procalcitonin  {
	di "`var'"

	nptrend `var' `flagD',gr(f_score) jterpstra
}

//Uses T16 after dropping (7) those cases identified by flagD in T15 for bing in the study before 11-30-19
//
//Table 5 data
```

```
tab any_lab

tab wbc_done

tab crp_done

tab esr_done

tab bnp_done

reg l_ldh  i.f_score i.age_group1  [pweight =iptwt_ldh_done ]

reg l_ldh  i.f_score  [pweight =iptwt_ldh_done ]

reg l_ldh  i.f_score i.b4.age_gr [pweight =iptwt_ldh_done ]

reg l_ldh  f4 i.b4.age_gr [pweight =iptwt_ldh_done ]

reg l_ldh  i.f_score i.b4.age_gr [pweight =iptwt_ldh_done ]

reg l_ferritin  i.f_score i.b4.age_gr [pweight =iptwt_ferritin_measure  ]

reg l_lft_ast   i.f_score i.b4.age_gr [pweight =iptwt_ast_done  ]

reg l_lft_alt   i.f_score i.b4.age_gr [pweight =iptwt_alt_done  ]

logistic  elev_dimer   i.f_score i.b4.age_gr [pweight =iptwt_dimer_measured  ]

logit elev_dimer f456 i.age_group [pw =iptwt_dimer_measured ]  ,or

        //

//

//

//

cap frame drop diagt_f456

frame create diagt_f456  str15(variable) double(ppv lb_ppv ub_ppv npv lb_npv ub_npv sens  lb_sens ub_sens spec
lb_spec ub_spec auc auc_lb auc_ub prev)

 foreach var in admit elev_ferritin elev_ldh elev_lft_alt elev_lft_ast elev_crp elev_esr elev_procalcitonin   elev_dimer
elev_trop {

        di "`var'"

        diagt `var' f456 if flagD==0,sf

        frame post diagt_f456   ("`var'") (r(ppv)) (r(ppv_lb)) (r(ppv_ub)) (r(npv)) (r(npv_lb)) (r(npv_ub)) ( r(sens))
(r(sens_lb)) (r(sens_ub)) (r(spec))  (r(spec_lb)) (r(spec_ub)) (r(roc))  (r(roc_lb))  (r(roc_ub)) (r(prev))
```

```
}

frame diagt_f456 :save diagt_f456 ,replace

cap frame drop diagt_f56

frame create diagt_f56  str15(variable) double(ppv lb_ppv ub_ppv npv lb_npv ub_npv sens  lb_sens ub_sens spec lb_spec ub_spec auc  auc_lb auc_ub prev)

 foreach var in admit elev_ferritin elev_ldh elev_lft_alt elev_lft_ast elev_crp elev_esr elev_procalcitonin   elev_dimer elev_trop {

	di "`var'"

	diagt `var' f56 if flagD==0,sf

	frame post diagt_f56   ("`var'") (r(ppv)) (r(ppv_lb)) (r(ppv_ub)) (r(npv)) (r(npv_lb)) (r(npv_ub)) ( r(sens)) (r(sens_lb)) (r(sens_ub)) (r(spec))  (r(spec_lb)) (r(spec_ub)) (r(roc)) (r(roc_lb))  (r(roc_ub))(r(prev))

}

frame diagt_f56 :save diagt_f56 ,replace
```

Appendix 4

Sample Ultrasound reports and classification.

Output is cleaned and de-identified but may include duplicates. (During the study period may reports were generated using templates which were modified for individual patients)

Format is:

“Start report“

“Report body”

“Interpretation”

Start report
"ultrasound  indication: rule out infiltrate/fluid  machine:  mindray   archived:  local   probe: 10.5 mhz  image quality:good  narrative: posterior , axillary and anterior acoustic windows interrogated bilaterally. normal a lines throughout. no excess b-lines, air bronchograms or effusion. no consolidation  impression:normal lung us "
"Normal"

Start report
"point of care lung ultrasound  indication: rule out infiltrate/fluid  machine:  mindray   archived:  local   probe: 10.5 mhz, 4.1 mhz  image quality: excellent  narrative: posterior , axillary and anterior acoustic windows interrogated bilaterally.   impression:patch of moth-eaten pleura and long b lines in the left infrascapular window. no effusion. no consolidation. "
"Very mild"

Start report
"ultrasound performed at bedside  point of care limited  lung ultrasound  indication: rule out infiltrate/fluid  machine: mindray   archived:  local   probe: 10.5 mhz  image quality: good   narrative: posterior , axillary and anterior acoustic windows interrogated bilaterally. excessive long b-lines with irregular air bronchiograms on the right. subpleural pneumonia, less than 1 cm on the right.   impression:abnormal
"Mild to Moderate"

Start report
"ultrasound  indication: rule out infiltrate/fluid  machine:  mindray   archived:  local   probe: 10.5 mhz  image quality: good  narrative: posterior , axillary and anterior acoustic windows interrogated bilaterally. normal a lines throughout. no excess b-lines, air bronchograms or effusion. no consolidation  impression:normal lung us
"Normal"

Start report
"ultrasound  bedside lung ultrasound shows small amount of patchy viral pneumonitis in the right axillary acoustic window and some mild pleural irregularities and the intrascapular windows.  images archived locally  machine: mindray  probe: 9–3 megahertz  impression: very mild pneumonitis     differential diagnoses are considered as follows, but not limited to: covid pneumonitis, other viral pneumonitis, pneumonia, adenovirus.

"Very mild"

Start report
point of care lung ultrasound  indication: rule out infiltrate/fluid  machine:  mindray   archived:  local   probe: 10.5 mhz  image quality: very good   narrative: posterior , axillar and anterior acoustic windows interrogated bilaterally. normal a lines, no excess b-lines, no effusion, no consolidation  impression: normal

"Normal"

Start report
"ultrasound. point of care limited lung ultrasound indication: rule out infiltrate/fluid machine: mindray archived: local probe: 4.1mhz image quality: good narrative: posterior , axillar and anterior acoustic windows interrogated bilaterally. impression: normal
"Normal"

Start report
"point of care lung ultrasound indication: rule out infiltrate/fluid machine: mindray archived: local probe: 10.5 mhz image quality: very good narrative: posterior , axillary and anterior acoustic windows interrogated bilaterally. air bronchograms. no excess b lines. normal short b lines. no consolidation. no effusion. impression:bronchitis "
"Very mild"

Start report
"ultrasound indication: rule out infiltrate/fluid machine: mindray archived: local probe: 10.5 mhz image quality: good narrative: posterior , axillary and anterior acoustic windows interrogated bilaterally. normal a lines throughout. no excess b-lines, air bronchograms or effusion. no consolidation impression:normal lung us "
"Normal"

Start report
"ultrasound radiology report sutter medical center emergency department - sacramento exam: poc lung ultrasound indications: known covid exposure with symptoms views: 2-3 views (as below) probe: linear probe image quality: good procedure: using the linear array transducer i evaluated the bilateral anterolateral thorax in a systematic fashion revealing the presence of lung sliding and comet-tail artifact at the interrogated interspaces. m-mode shows normal anatomy and function. impression: negative limited lung ultrasound with no pneumothorax identified. these images were archived and i independently interpreted the images at the bedside.
"Normal"

Start report
"ultrasound indication: rule out infiltrate/fluid machine: mindray archived: local probe: 10.5 mhz image quality: good narrative: posterior , axillary and anterior acoustic windows interrogated bilaterally. impression: normal lung exam
"Normal"

Start report
" point of care limited lung ultrasound indication: rule out infiltrate/fluid machine: mindray archived: local probe: 10.5 mhz image quality: good narrative: posterior , axillary and anterior acoustic windows interrogated bilaterally. excess long b lines right intrascapular area. some moth eaten pleura bilat axillae clear. no effusion or consolidation impression:pneumonitis c/w covid
"Mild"

Start report
" procedures (if indicated) .ed ultrasound indications: rule out pneumonia imaged archived locally in ultrasound machine. a 10 mghz ultrasound used to interrogate lung fields and heart fields. on us, patient has excess b lines with areas of consolidation, as well as pleural thickening of the right lung. there are shorter b lines notable in the

left upper lung. there were no signs of cardiac effusions. there is normal contractivity. "
"Moderate"

Start report
"ultrasound: lung indications: rule out pneumonia imaged archived locally in mindray ltrasound machine. a 10 mghz ultrasound used to interrogate all posterior lung fields and long and short parasternal axis cardiac fields. on us, patient has excess long b lines bilaterally with areas of consolidation, as well as pleural thickening of the right lung. there are excess short b lines without excess long notable in the both upper lungs. there was no pericardial effusions. there is normal contractility of heart. impression: bilateral alveloar fluid and patchy consolidation on the right with small amount of pleural effuison "
"Moderate"

Start report
"limited bedside poc ultrasound radiology report sutter medical center emergency department - sacramento exam: poc lung ultrasound indications: chest pain or shortness of breath views: 2-3 views (as below) probe: linear probe image quality: good procedure: using the linear array transducer i evaluated the bilateral posterolateral thorax in a systematic fashion revealing the presence of lung sliding with pleural thickening irregularities and b lines in some of the lung fields. impression: findings consistent with covid. these images were archived and i independently interpreted the images at the bedside.
"Mild"

Start report
"ultrasound indication: rule out infiltrate/fluid machine: mindray archived: local probe: 4-1mhz image quality: good narrative: posterior , axillary and anterior acoustic windows interrogated bilaterally. normal a lines throughout. no excess b-lines, air bronchograms or effusion. no consolidation. one area of moth eaten pleura. impression:normal lung us this document was transcribed by nicolo "
"Very mild"

Start report
" bedside ultrasound showed no evidence of pleural thickening or irregularities. she had no evidence of consolidations or b lines on ultrasound."
"Normal"

Start report
" point of care lung ultrasound indication: rule out infiltrate/fluid machine: mindray archived: local probe: 10-.5 mhz image quality: good narrative: posterior , axillar and anterior acoustic windows interrogated bilaterally. upper limitof normal short b lines. normal a lines. no effusion"
"Normal"

Start report
" point of care limited lung ultrasound indication: rule out infiltrate/fluid machine: mindray archived: local probe: 10.5 mhz image quality: good narrative: posterior , axillary and anterior acoustic windows interrogated bilaterally. impression:normal cardiac windows with no effusion.
"Normal"

Start report
"   point of care limited lung ultrasound     indication: rule out infiltrate/fluid  machine:  mindray   archived:  local   probe: 9-3mhz  image quality: good   narrative: posterior , axillary and anterior acoustic windows interrogated bilaterally. normal a lines throughout. no excess b-lines, air bronchograms or effusion.  "
"Normal"

Start report
"          point of care lung ultrasound  indication: rule out infiltrate/fluid  machine:  mindray   archived:  local   probe: 10-5 mhz  image quality: good  narrative: posterior , axillary and anterior acoustic windows interrogated bilaterally. she has normal a lines, no excess b-lines, and no consolidation or effusion.  impression: normal
"Normal"

Start report
"point of care lung ultrasound  indication: rule out infiltrate/fluid  machine:  mindray   archived:  local   probe: 4.1mhz image quality: fair   narrative: posterior , axillar and anterior acoustic windows interrogated bilaterally. moth-eaten pleura in and excess long coalescent b lines  in right > left intrascaupluar windows and  axillary windows. no effusion or focal consolidation     impression: pneumonitis c/w covid
"Mild to Moderate"

Start report
"point of care lung ultrasound  indication: rule out infiltrate/fluid  machine:  mindray   archived:  local   probe: 10.5 mhz  image quality: ***  narrative: posterior acoustic windows interrogated bilaterally.   impression:borderline excess B lines in 1 field.   "
"Very mild"

Start report
"point of care lung ultrasound  indication: rule out infiltrate/fluid  machine:  mindray   archived:  local   probe: 10.5 mhz  image quality: normal  narrative: posterior , axillary and anterior acoustic windows interrogated bilaterally. normal  impression: normal    "
"Normal"

Start report
"    point of care lung ultrasound  indication: rule out infiltrate/fluid  machine:  mindray   archived:  local   probe: 10.5 mhz  image quality: very good   narrative: posterior , axillar and anterior acoustic windows interrogated bilaterally. normal a lines,. upper liit normal short b lines. no long b lines no effusion or consolidation   impression:normal     "
"Normal"

Start report
"ultrasound  indication: rule out infiltrate/fluid  machine:  mindray   archived:  local   probe: 10.5 mhz  image quality: good  narrative: posterior , axillary and anterior acoustic windows interrogated bilaterally. normal a lines throughout. no excess b-lines, air bronchograms or effusion. no consolidation  impression:normal lung us
"Normal"

Start report
"ultrasound was done showing a subcentimeter consol"
"Very mild"

Start report
" limited bedside poc ultrasound radiology report sutter medical center emergency department - sacramento exam: poc limited lung ultrasound indications: chest pain views: 2-3 views (as below) probe: linear array image quality: excellent procedure: using the linear array transducer i evaluated the bilateral anterior, posterior, superior and inferior thorax in a systematic fashion "
"Normal"

Start report
"ultrasound. consistent with very mild covid. see report below. point of care limited lung ultrasound indication: rule out infiltrate/fluid machine: mindray archived: local probe: 9-3 mhz image quality: good narrative: posterior , axillary and anterior acoustic windows interrogated bilaterally. pleural thickening intrascapular r>l. no excessive b lines. consistent with very mild covid. impression: normal lung us
"Very mild"

Start report
") us: point of care lung ultrasound indication: rule out infiltrate/fluid machine: mindray archived: local probe: 10.5 mhz image quality: *** narrative: posterior , axillar and anterior acoustic windows interrogated bilaterally. excess b lines. excess air bronchograms. crowded, consistent with right base. lower acoustic window and right axilla. upper limit of normal. short b lines. impression: lobar pneumonia on right. medical decision making "
"Moderate"

Start report
"ultrasound indication: rule out infiltrate/fluid machine: mindray archived: local probe: 10.5 mhz image quality: very good narrative: posterior , axillary and anterior acoustic windows interrogated bilaterally. normal a lines throughout. no excess b-lines, air bronchograms or effusion. no consolidation . one inch spot with increased long b line in the right intrascapular regioun impression:normal lung us
"Very mild"

Start report
" ultrasound probe 1 impressions: normal. no effusions. no b lines. normal a lines. **PM patient seen and examined. ultrasound is reassuring. will order covid swab. advised mother of return precautions."
"Normal"

Start report
"ultrasound indication: rule out infiltrate/fluid machine: mindray archived: local probe: 10.5 mhz image quality: good narrative: posterior , axillary and anterior acoustic windows interrogated bilaterally. normal a lines throughout. no excess b-lines, air bronchograms or effusion. no consolidation impression:normal lung us
"Normal"

Start report
"ultrasound  indication: rule out infiltrate/fluid  machine:  mindray   archived:  local   probe: 10.5 mhz  image quality: good  narrative: posterior , axillary and anterior acoustic windows interrogated bilaterally. normal a lines throughout. no excess b-lines, air bronchograms or effusion. no consolidation  impression:normal lung us "
"Normal"

Start report
"ultrasound  indication: rule out infiltrate/fluid  machine:  mindray   archived:  local   probe: 10.5 mhz  image quality: normal  narrative: posterior , axillary and anterior acoustic windows interrogated bilaterally. normal a lines throughout. no excess b-lines, air bronchograms or effusion. no consolidation. small area with slightly rough pleura impression:normal lung us
"Very mild"

Start report
"ultrasound  indication: rule out infiltrate/fluid  machine:  mindray   archived:  local   probe: 10.5 mhz  image quality: good  narrative: posterior , axillary and anterior acoustic windows interrogated bilaterally. normal a lines throughout. no excess b-lines, air bronchograms or effusion. no consolidation  impression:normal lung us
"Normal"

Start report
"ultrasound radiology report  sutter medical center emergency department - sacramento    exam: poc lung ultrasound indications: known covid exposure with symptoms    views: 2-3 views (as below)    probe: linear probe    image quality: good    procedure: using the linear array transducer i evaluated the bilateral anterolateral thorax in a systematic fashion revealing the presence of lung sliding and comet-tail artifact at the interrogated interspaces. m-mode shows normal anatomy and function.     impression: negative limited lung ultrasound with no pneumothorax identified.  these images were archived and i independently interpreted the images at the bedside.
"Normal"

Start report
"  point of care lung ultrasound  indication: rule out infiltrate/fluid  machine:  mindray   archived:  local   probe: 10.5 mhz  image quality: good  narrative: posterior , axillar and anterior acoustic windows interrogated bilaterally.significant excess short and long b lines intrascapular region. some pleura thickening. no effusion or consolidation "
"Mild to Moderate"

Start report
"point of care lung ultrasound  indication: rule out infiltrate/fluid  machine:  mindray   archived:  local   probe: 10.5 mhz  image quality:good   narrative: posterior , axillar and anterior acoustic windows interrogated bilaterally. excess intrascapular  b lines. no effusion or consolidation.   impression:pneumonitis c/w covid      "
"Mild"

Start report
"  point of care limited  lung ultrasound  indication: rule out infiltrate/fluid  machine:  mindray   archived:  local   probe: 10.5 mhz  image quality: good   narrative: posterior , axillary and anterior acoustic windows interrogated bilaterally. few air bronchograms. no excess b lines. no effusion. no consolidation.  impression c/w mild bronchitis. no

evidence for covid-19. "
"Very mild"

Start report
"ultrasound indication: rule out infiltrate/fluid machine: mindray archived: local probe: 9-3 mhz image quality: good narrative: posterior , axillary and anterior acoustic windows interrogated bilaterally. he has some moth-eaten pleura and occasional excess long b-lines in the left and right axilla. the posterior and anterior windows are normal. no effusion. no consolidation. impression: abnormal lung us consistent with very mild pneumonitis. very mild viral pneumonitis, left and right axillae. "
"Very mild"

Start report
" bedside ultrasound performed showing no signs of covid-19. "
"Normal"

Start report
" point of care lung ultrasound indication: rule out infiltrate/fluid machine: mindray archived: local probe: 10.5 mhz image quality: good narrative: posterior acoustic windows interrogated bilaterally. normal a lines. occasional b lines in normal range. no pleural effusion. impression: *** "
"Normal"

Start report
"ultrasound indication: rule out infiltrate/fluid machine: mindray archived: local probe: 9-3mhz image quality: good narrative: posterior , axillary and anterior acoustic windows interrogated bilaterally. normal a lines throughout. no excess b-lines, air bronchograms or effusion. no consolidation impression:normal lung us
"Normal"

Start report
" point of care lung ultrasound indication: rule out infiltrate/fluid machine: mindray archived: local probe: 4.1mhz image quality: good narrative: posterior , axillar and anterior acoustic windows interrogated bilaterally. *** impression:*** "
"Normal"

Start report
"ultrasound indication: rule out infiltrate/fluid machine: mindray archived: local probe: 10.5 mhz image quality: good narrative: posterior , axillary and anterior acoustic windows interrogated bilaterally. normal a lines throughout. no excess b-lines, air bronchograms or effusion. no consolidation impression:normal lung us
"Normal"

Start report
" indication: rule out infiltrate/fluid machine: mindray archived: local probe: 9-3 mhz image quality: very goood narrative: posterior , axillary and anterior acoustic windows interrogated bilaterally. there are extensive b-lines and associated moth eaten pleura intrascapularly left greater than right and in the axilla to a lesser extent. impression: abnormal lung us consistent with covid. "
"Moderate to severe"

Start report
"ultrasound  indication: rule out infiltrate/fluid  machine:  mindray   archived:  local   probe: 10.5 mhz  image quality: goodnarrative: posterior , axillary and anterior acoustic windows interrogated bilaterally. normal a lines throughout. no excess b-lines, air bronchograms or effusion. no consolidation  impression:normal lung us
"Normal"

Start report
"ultrasound   indication: rule out infiltrate/fluid  machine:  mindray   archived:  local   probe: 9-3 mhz  image quality: good   narrative: posterior , axillary and anterior acoustic windows interrogated bilaterally. moth-eaten pleura bilaterally. no effusion or consolidation. compression consistent with very mild viral pneumonitis.  impression: abnormal lung us     "
"Mild"

Start report
"  point of care lung ultrasound  indication: rule out infiltrate/fluid  machine:  mindray   archived:  local   probe: 10.5 mhz  image quality: good  narrative: posterior and axillar acoustic windows interrogated bilaterally.   impression: diffuse a-lines, no evidence of b-lines, no effusion  "
"Normal"

Start report
" point of care lung ultrasound  indication: rule out infiltrate/fluid  machine:  mindray   archived:  local   probe: 10.5 mhz  image quality: good  narrative: posterior and axillar acoustic windows interrogated bilaterally.   impression: diffuse a-lines, no evidence of b-lines, no effusion"
"Normal"

Start report
"ultrasound  indication: rule out infiltrate/fluid  machine:  mindray   archived:  local   probe: 10.5 mhz  image quality: good  narrative: posterior , axillary and anterior acoustic windows interrogated bilaterally. normal a lines throughout. no excess b-lines, air bronchograms or effusion. no consolidation  impression:normal lung us"
"Normal"

Start report
"   point of care lung ultrasound  indication: rule out infiltrate/fluid  machine:  mindray   archived:  local   probe: 10.5 mhz  image quality: goodnarrative: posterior , axillary and anterior acoustic windows interrogated bilaterally. 1 right sided subpleural subcentimeter focal pneumonia. no excess b-lines elsewhere. no effusion. no significant consolidation. impression: as this finding was isolated no treatment indicated.   "
"Very mild"
"

Start report
"ultrasound     indication: rule out infiltrate/fluid  machine:  mindray   archived:  local   probe: 9-3mhz  image quality: good   narrative: posterior , axillary and anterior acoustic windows interrogated bilaterally. normal a lines throughout. no excess b-lines, air bronchograms or effusion. no consolidation  impression:normal lung us        reviewed and electronically signed by paul walsh md.
"Normal"

Start report
"  point of care limited  lung ultrasound  indication: rule out infiltrate/fluid  machine:  mindray   archived:  local probe: 10.5 mhz  image quality: good   narrative: posterior , axillary and anterior acoustic windows interrogated bilaterally. diffuse excess long coalescent b lines. bronchograms in the left. no pleural effusion or consolidation. impression: findings consistent with moderate severity pneumonitis, bronchiolitis or chf.
"Moderate"

Start report
"ultrasound  indication: rule out infiltrate/fluid  machine:  mindray   archived:  local   probe: 10.5 mhz  image quality: good  narrative: posterior , axillary and anterior acoustic windows interrogated bilaterally. normal a lines throughout. no excess b-lines, air bronchograms or effusion. no consolidation  impression:normal lung us
"Normal"

Start report
" the ultrasound doesn't show any evidence of lung infection."
"Normal"

Start report
"  point of care lung ultrasound  indication: rule out infiltrate/fluid  machine:  mindray   archived:  local   probe: 10.5 mhz  image quality: very good.  narrative: posterior , axillar and anterior acoustic windows interrogated bilaterally. normal a-lines. no b-lines.  impression: no effusion. no consolidation.  "
"Normal"

Start report
"  point of care lung ultrasound  indication: rule out infiltrate/fluid  machine:  mindray   archived:  local   probe: 10.5 mhz  image quality: very good.  narrative: posterior , axillary and anterior acoustic windows interrogated bilaterally. normal a-lines. no b-lines. no effusion. no consolidation.    impression:normal    this document was transcribed by julianna c rojo, cms.    signature & attestation: all medical record entries made
"Normal"

Start report
"ultrasound  indication: rule out infiltrate/fluid  machine:  mindray   archived:  local   probe: 10.5 mhz  image quality: good   narrative: posterior , axillary and anterior acoustic windows interrogated bilaterally. excess b lines in the right posterior lung field excess b lines in the right posterior lung. some moth-eaten pleura i nthe left axilla.   impression: mild patchy pneumonitis c/w covid     "
"Mild"

Start report
"point of care lung ultrasound  indication: rule out infiltrate/fluid  machine:  mindray   archived:  local   probe: 10.5 mhz  image quality: normal  narrative: posterior , axillary and anterior acoustic windows interrogated bilaterally. normal  impression:normal     "
"Normal"

Start report
"point of care lung ultrasound  indication: rule out infiltrate/fluid  machine:  mindray   archived:  local   probe: 10.5 mhz  image quality: normal  narrative: posterior , axillary and anterior acoustic windows interrogated bilaterally. normal  impression:normal     "
"Normal"

Start report
"ultrasound   indication: rule out infiltrate/fluid  machine:  mindray   archived:  local   probe: 9-3 mhz  image quality: good   narrative: posterior , axillary and anterior acoustic windows interrogated bilaterally. moth-eaten pleura in the intrascapular windows, occasional b-lines, no effusion or consolidation   impression: abnormal lung us, consistent with mild viral pneumonitis  "
"Mild"

Start report
"  point of care lung ultrasound  indication: rule out infiltrate/fluid  machine:  mindray   archived:  local   probe: 10.5 mhz and 4.1 mhz  image quality: fair   narrative: posterior acoustic windows interrogated bilaterally. excess b lines intrascapular bilat r >left with  localized consolidation on the right. no effusion + air bronchograms right. no moth eaten pleura.  impression: consolidation in the right intrascapular area with excess b-lines consistent with covid or other patchy pneum"
"Mild to Moderate"

Start report
"ultrasound   indication: rule out infiltrate/fluid  machine:  mindray   archived:  local   probe: 9-3 mhz  image quality: good   narrative: posterior , axillary and anterior acoustic windows interrogated bilaterally.  there were multiple areas of long excess b-lines mostly in the right interscapular area with less severe findings in the left interscapular area and relative sparing of the axilla.  no effusion no focal consolidations  impression: abnormal lung us consistent with viral pneumonitis.
"Moderate to severe"

Start report
"ultrasound  indication: rule out infiltrate/fluid  machine:  mindray   archived:  local   probe: 10.5 mhz  image quality: goodd  narrative: posterior , axillary and anterior acoustic windows interrogated bilaterally. normal a lines throughout. no excess b-lines, air bronchograms or effusion. no consolidation. one window less than upper limit of b lines only. cardiac window showed normal contractility   impression:normal lung us
"Normal"

Start report
"   point of care limited lung ultrasound   indication: rule out infiltrate/fluid  machine:  mindray   archived:  local probe: 9-3 mhz  image quality: good   narrative: posterior , axillary and anterior acoustic windows interrogated bilaterally.  he has long b-lines with moth-eaten pleura right posterior acoustic windows, left posterior acoustic windows and right axilla.  he has several pustule is less than 0.5 cm each with some surrounding irregular air bronchograms "
"Moderate"

Start report
"    point of care lung ultrasound  indication: rule out infiltrate/fluid  machine:  mindray   archived:  local   probe: 10.5 mhz  image quality: good  narrative: posterior , axillary and anterior acoustic windows interrogated bilaterally. impression:  excess short B lines diffusely on the posterior windows. moth eaten pleural diffusely bilaterally on the left lower bases. excess long B lines in the left axilla. findings are consistent with viral pneumonitis or pneumonia.   "
"Moderate"

Start report
"point of care lung ultrasound  indication: rule out infiltrate/fluid  machine:  mindray   archived:  local   probe: 10.5 mhz  image quality: 1  narrative: posterior , axillary and anterior acoustic windows interrogated bilaterally. the left has some increased b lines on the right base. excess long b lines.  impression: patchy viral pneumonitis"
"Mild"

Start report
"ultrasound     indication: rule out infiltrate/fluid  machine:  mindray   archived:  local   probe: 9-3mhz  image quality: good   narrative: posterior , axillary and anterior acoustic windows interrogated bilaterally. normal a lines throughout. no excess b-lines, air bronchograms or effusion. no consolidation  impression:normal lung us    .    "Normal"

Start report
"ultrasound  indication: rule out infiltrate/fluid  machine:  mindray   archived:  local   probe: 10.5 mhz  image quality: good  narrative: posterior , axillary and anterior acoustic windows interrogated bilaterally. normal a lines throughout. minimal b-lines left lung with no air bronchograms or effusion. no consolidation  impression:normal lung us   "
"Very mild"

Start report
"  point of care limited  lung ultrasound  indication: rule out infiltrate/fluid  machine:  mindray   archived:  local   probe: 10.3 mhz  image quality: good   narrative: posterior , axillary and anterior acoustic windows interrogated bilaterally. ***  findings: moth eaten plethora in bilateral intrascapular and acoustic window.   impression: mild penumonitis        "
"Mild"

Start report
"  point of care limited  lung ultrasound  indication: rule out infiltrate/fluid  machine:  mindray   archived:  local   probe: 10.3 mhz  image quality: good   narrative: posterior , axillary and anterior acoustic windows interrogated bilaterally.    findings: moth eaten plethora in bilateral intrascapular and acoustic window. no effusion or consolidation impression: mild penumonitis      "
"Mild"

Start report
"  point of care limited  lung ultrasound  indication: rule out infiltrate/fluid  machine:  mindray   archived:  local   probe: 10.5 mhz  image quality: good   narrative: posterior , axillary and anterior acoustic windows interrogated bilaterally. left upper lobe with slight increase in b lines that are mostly short. on the right axilla , excess long b lines.

impression very mild patchy pneumonitis "
"Very mild"

Start report
" point of care lung ultrasound indication: rule out infiltrate/fluid machine: mindray archived: local probe: 10.5 mhz image quality: good narrative: posterior , axillary and anterior acoustic windows interrogated bilaterally with evidence of scattered b lines. impression: scattered b lines "
"Very mild"

Start report
" point of care limited lung ultrasound indication: rule out infiltrate/fluid machine: mindray archived: local probe: 10.5 mhz image quality: good narrative: posterior , axillary and anterior acoustic windows interrogated bilaterally. *** impression: left lung with multiple long b lines, mostly linear bronchoairgrams. left lung base there are areas of multifocal irregularly placed air bronchograms c/w pneumonia. tiny pleural effusion on the right base with irregular areas of consolidation about 1 cm depth. "
"Moderate"

Start report
"point of care limited lung ultrasound indication: rule out infiltrate/fluid machine: mindray archived: local probe: 10.5 mhz image quality: good narrative: posterior , axillary and anterior acoustic windows interrogated bilaterally. right lung: with multiple long b lines , beneath this linear bronchoairgrams. beneath this near the right lung base there are areas of multifocal irregularly placed air bronchograms c/w pneumonia. left lung tiny pleural effusion on the left base with irregular areas of consolidation about 1 cm depth. impression :extensive right sided consolidation c/w pna and smuch smallef left sided pna "
"Severe"

Start report
"ultrasound indication: rule out infiltrate/fluid machine: mindray archived: local probe: 9-3 mhz image quality: good narrative: posterior , axillary and anterior acoustic windows interrogated bilaterally. diffuse air bronchogram's on left lung with b lines in base. irregularly spaced air bronchograms and pleural thickening. b lines on left base and left midzone. depth greater than 1 cm. long b lines in upper posterior acoustic constrict windowns. some b lines located on right upper. irregularly spaced air bronchogram's on right base consistent with pneumonia. incidental finding of mild hydronephrosis on left kidney. impression: abnormal lung us "
"Moderate"

Start report
"ultrasound radiology report sutter medical center emergency department - sacramento exam: poc limited lung ultrasound indications: fever or shortness of breath views: 2-3 views (as below) probe: linear array image quality: good procedure: using the linear array transducer i evaluated the bilateral anterior, posterior and lateral thorax in a systematic fashion revealing the presence of significant consolidation lll. mild diffuse b lines. there is the presence of lung sliding and comet-tail artifact at the interrogated interspaces. m-mode shows normal anatomy and function. impression: point of care limited pediatric lung ultrasound with the presence of a consolidation consistent with pneumonia absence of a pneumonthorax and findings consistent with bronchiolitis. "

"Mild to Moderate"

Start report
"ultrasound indication: rule out infiltrate/fluid machine: mindray archived: local probe: 9-3 mhz image quality: good narrative: posterior , axillary and anterior acoustic windows interrogated bilaterally. excess long b lines and moth eaten pluera at left base and right area with no focal consolictiaon impression: consistent with covid, abnormal lung us "
"Mild to Moderate"

Start report
"ultrasound indication: rule out infiltrate/fluid machine: mindray archived: local probe: 9-3mhz image quality: good narrative: posterior , axillary and anterior acoustic windows interrogated bilaterally. normal a lines throughout. no excess b-lines, air bronchograms or effusion. no consolidation impression:normal lung us "
"Normal"

Start report
"ultrasound. indication: rule out infiltrate/fluid machine: mindray archived: local probe: 9-3 mhz image quality: very good narrative: posterior , axillary and anterior acoustic windows interrogated bilaterally. diffuse excess long b lines, moth-eaten pleura. consistent with covid pneumonitis. impression: abnormal lung us
"Severe"

Start report
"point of care lung ultrasound indication: rule out infiltrate/fluid machine: mindray archived: local probe: 10.5 mhz image quality: good narrative: posterior , axillary and anterior acoustic windows interrogated bilaterally he has 2 tiny foci of excess long b-lines one in the left axilla and one in the right intrascapular posterior window. impression: consistent with very mild covid pneumonitis records reviewed: nursing notes relevant past records "
"Very mild"

Start report
"ultrasound indication: rule out infiltrate/fluid machine: mindray archived: local probe: 10.5 mhz image quality:good narrative: posterior , axillary and anterior acoustic windows interrogated bilaterally. normal a lines throughout. no excess b-lines, air bronchograms or effusion. no consolidation impression:normal lung us
"Normal"

Start report
"ultrasound indication: rule out infiltrate/fluid machine: mindray archived: local probe: 10.5 mhz image quality: good narrative: posterior , axillary and anterior acoustic windows interrogated bilaterally. normal a lines throughout. no excess b-lines, air bronchograms or effusion. no consolidation impression:normal lung us "
"Normal"

Start report
"ultrasound indication: rule out infiltrate/fluid machine: mindray archived: local probe: 10.5 mhz image quality: good narrative: posterior , axillary and anterior acoustic windows interrogated bilaterally. normal a lines throughout. no excess b-lines, air bronchograms or effusion. no consolidation impression:normal lung us "
"Normal"

Start report
"ultrasound sutter medical center emergency department - sacramento exam: poc lung ultrasound indications: rule out pneumonia probe: 10.5 mghz linear probe image quality: good procedure: using the linear array transducer i evaluated the bilateral anterolateral thorax in a systematic fashion revealing the presence of lung sliding and comet-tail artifact at the interrogated interspaces. m-mode shows normal anatomy and function. findings: excess long and short B lines throughout right lung. B lines in left lung field is the upper limit of normal. impression:findings suggest pneumonia in the right lung field. left lung fields are negative for pneumonia. these images were archived and i in"
"Mild to Moderate"

Start report
"ultrasound indication: rule out infiltrate/fluid machine: mindray archived: local probe: 9-3mhz image quality: poor because child unable to sit still. narrative: posterior , axillary and anterior acoustic windows interrogated bilaterally. normal a lines throughout. no excess b-lines, air bronchograms or effusion. no consolidation impression:normal lung us "
"Normal"

Start report
" point of care limited lung ultrasound indication: rule out infiltrate/fluid machine: mindray archived: local probe: 10.5 mhz image quality: good narrative: posterior , some upper limit of normal long b lines shown in right axilla. left axilla normal. impression: consistent with early pneumonitis but very mild "
"Very mild"

Start report
"ultrasound indication: rule out infiltrate/fluid machine: mindray archived: local probe: 10.5 mhz image quality: good narrative: posterior , axillary and anterior acoustic windows interrogated bilaterally. normal a lines throughout. no excess b-lines, air bronchograms or effusion. no consolidation impression:normal lung us "
"Normal"

Start report
"1 point of care limited lung ultrasound indication: rule out infiltrate/fluid machine: mindray archived: local probe: 10.5 mhz image quality: good narrative: posterior , axillary and anterior acoustic windows interrogated bilaterally. some minor moth eaten pleura and long b lines intrasacpularly bilaterally. very mild no axillary involvement. no effusion/consolidation impression:borderline for very mild pneumonitis
"Very mild"

Start report
"1point of care limited lung ultrasound indication: rule out infiltrate/fluid machine: mindray archived: local probe: 10.5 mhz image quality: good narrative: posterior , axillary and anterior acoustic windows interrogated bilaterally. some minor moth eaten pleura and long b lines intrasacpularly bilaterally. very mild no axillary involvement. no effusion/consolidation impression:borderline for very mild pneumonitis "
"Very mild"

Start report
"ultrasound indication: rule out infiltrate/fluid machine: mindray archived: local probe: 10.5 mhz image quality: good narrative: posterior , axillary and anterior acoustic windows interrogated bilaterally. occasional air bronchograms. impression: essentially normal lung ultrasound with tiny amount of bronchitis
"Very mild"

Start report
" ultrasound probe 1 impression: normal. no B lines. normal a lines. no pleural thickening. no effusions or consolidations. ultrasound is normal. "
"Normal"

Start report
"ultrasound indication: rule out infiltrate/fluid machine: mindray archived: local probe: 9-3mhz image quality: good narrative: posterior , axillary and anterior acoustic windows interrogated bilaterally. normal a lines throughout. no excess b-lines, air bronchograms or effusion. no consolidation impression:normal lung us
"Normal"

Start report
"ultrasound indication: rule out infiltrate/fluid machine: mindray archived: local probe: 10.5 mhz image quality: good narrative: posterior , axillary and anterior acoustic windows interrogated bilaterally. normal a lines throughout. no excess b-lines, air bronchograms or effusion. no consolidation impression:normal lung us
"Normal"

Start report
" point of care limited lung ultrasound indication: rule out infiltrate/fluid machine: mindray archived: local probe: 10.5 mhz image quality: good narrative: posterior , axillary and anterior acoustic windows interrogated bilaterally. mild and long b-lines with mild moth-eaten pleura. no effusion no consolidation. impression: consistent with mild pneumonitis. "
"Mild"

Start report
"ultrasound indication: rule out infiltrate/fluid machine: mindray archived: local probe: 10.5 mhz image quality: good narrative: posterior , axillary and anterior acoustic windows interrogated bilaterally. increased linear air bronchiograms bilaterally. no effusion or consolidation. impression:abnormal "
"Mild"

Start report
"ultrasound indication: rule out infiltrate/fluid machine: mindray archived: local probe: 9-3 mhz image quality: good narrative: posterior , axillary and anterior acoustic windows interrogated bilaterally. she has bilateral moth-eaten pleura but only rare b-lines. sparing of the axilla. no effusion, no consolidation impression: abnormal lung us consistent with very mild pneumonitis "
"Very mild"

Start report
"ultrasound indication: rule out infiltrate/fluid machine: mindray archived: local probe: 10.5 mhz image quality: normal narrative: posterior , axillary and anterior acoustic windows interrogated bilaterally. normal a lines throughout. no excess b-lines, air bronchograms or effusion. no consolidation impression:normal lung us medical decision making "
"Normal"

Start report
"ultrasound indication: rule out infiltrate/fluid machine: mindray archived: local probe: 10.5 mhz image quality: good narrative: posterior , axillar and anterior acoustic windows interrogated bilaterally. normal A lines. normal B lines. excess B lines with pleural thickening in left axillary window . no effusion. impression: pneumonia
"Mild to Moderate"

Start report
"limited bedside poc ultrasound radiology report sutter medical center emergency department - sacramento exam: poc limited lung ultrasound indications: fever or shortness of breath views: 2-3 views (as below) probe: linear array image quality: good procedure: using the linear array transducer i evaluated the bilateral anterior, posterior and lateral thorax in a systematic fashion revealing the presence of scatteredt b-lines there is the presence of lung sliding and comet-tail artifact at the interrogated interspaces. impression: point of care limited pediatric lung ultrasound with the absence of a consolidation consistent with pneumonia absence of a pneumonthorax and findings consistent with bronchiolitis. these images were archived and i independently interpreted the images at the bedside. "
"Mild"

Start report
"ultrasound indication: rule out infiltrate/fluid machine: mindray archived: local probe: 4-1mhz image quality: good narrative: posterior , axillary and anterior acoustic windows interrogated bilaterally. normal a lines throughout. no excess b-lines, air bronchograms or effusion. no consolidation impression:normal lung us. short b-lines in axilla.
"
"Normal"

Start report
" point of care lung ultrasound indication: rule out infiltrate/fluid machine: mindray archived: local probe: 10.5 mhz image quality: good narrative: posterior and axillar acoustic windows interrogated bilaterally. impression: diffuse a-lines, no evidence of b-lines, no effusion "
"Normal"

Start report
"ultrasound indication: rule out infiltrate/fluid machine: mindray archived: local probe: 10.5 mhz image quality: good narrative: posterior , axillary and anterior acoustic windows interrogated bilaterally. normal a lines throughout. few normal short b lines no excess b-lines, air bronchograms or effusion. no consolidation impression:normal lung us
"Normal"

Start report
"point of care lung ultrasound indication: rule out infiltrate/fluid machine: mindray archived: local probe: 10.5 mhz image quality: excellent narrative: posterior , axillar and anterior acoustic windows interrogated bilaterally. impression: normal. normal a lines. no b lines. no consolidation.
"Normal"

Start report
"ultrasound indication: rule out infiltrate/fluid machine: mindray archived: local probe: 10.5 mhz image quality: good narrative: posterior , axillary and anterior acoustic windows interrogated bilaterally. normal a lines throughout. no excess b-lines, air bronchograms or effusion. no consolidation impression:normal lung us upper limit of normal short b-lines diffusely. "
"Normal"

Start report
"ultrasound indication: rule out infiltrate/fluid machine: mindray archived: local probe: 10.5 mhz image quality: good narrative: posterior , axillary and anterior acoustic windows interrogated bilaterally. normal a lines throughout. no excess b-lines, air bronchograms or effusion. no consolidation impression:normal lung us reviewed and electronically signed by paul walsh md. "
"Normal"

Start report
" ultrasound was unremarkable. ","Normal"

161
"ultrasound radiology report sutter medical center emergency department - sacramento exam: poc lung ultrasound indications: chest pain or shortness of breath views: 2-3 views (as below) probe: linear probe image quality: good procedure: using the linear array transducer i evaluated the bilateral anterolateral thorax in a systematic fashion revealing the presence of lung sliding and comet-tail artifact at the interrogated interspaces. m-mode shows normal anatomy and function. impression: negative limited lung ultrasound with no pneumothorax ident"
"Normal"

Start report
"ultrasound indication: rule out infiltrate/fluid machine: mindray archived: local probe: 4.1mhz image quality: narrative: posterior , axillary acoustic windows interrogated bilaterally. he has excess b-lines with pleural thickening in the intrascapular acoustic windows bilaterally and axilla bilaterally. impression viral pneumonitis versus bronchiolitis "
"Moderate"

Start report
"ultrasound indication: rule out infiltrate/fluid machine: mindray archived: local probe: 10.5 mhz image quality:good narrative: posterior , axillary and anterior acoustic windows interrogated bilaterally. normal a lines throughout. no excess b-lines, air bronchograms or effusion. no consolidation impression:normal lung us "
"Very mild"

Start report
"ultrasound  indication: rule out infiltrate/fluid  machine:  mindray   archived:  local   probe: 10.5 mhz  image quality: good  narrative: posterior , axillary and anterior acoustic windows interrogated bilaterally. normal a lines throughout. no excess b-lines, air bronchograms or effusion. no consolidation  impression:normal lung us   "
"Normal"

Start report
"    point of care lung ultrasound  indication: rule out infiltrate/fluid  machine:  mindray   archived:  local   probe: 10.5 mhz  image quality: good  narrative: posterior , axillar and anterior acoustic windows interrogated bilaterally. no consolidation. normal a-line normal b lines   impression:normal    "
"Normal"

Start report
"ultrasound     indication: rule out infiltrate/fluid  machine:  mindray   archived:  local   probe: 9-3mhz  image quality: good   narrative: posterior , axillary and anterior acoustic windows interrogated bilaterally. normal a lines throughout. no excess b-lines, air bronchograms or effusion. no consolidation  impression:normal lung us        reviewed and electronically signed by paul walsh md.
"Normal"

Start report
" point of care lung ultrasound  indication: rule out infiltrate/fluid  machine:  mindray   archived:  local   probe: 10.5 mhz  image quality: good   narrative: posterior , axillary and anterior acoustic windows interrogated bilaterally.  he has excess long and short b-lines bilaterally in the intrascapular windows and in the right axilla. no effusion. some air bronchograms. no consolidation.  impression: ultrasound consistent with pneumonitis/bronchiolitis.     "
"Mild to Moderate"

Start report
"    point of care lung ultrasound  indication: rule out infiltrate/fluid  machine:  mindray   archived:  local   probe: 10.5 mhz  image quality: very good   narrative: posterior , axillar and anterior acoustic windows interrogated bilaterally. excess short b lines diffusely. thickened pleura. long blines upper limit normal - occassionally beyond that in some windows. no effusion   impression: pneumonitis - c/w covid"
"Mild"

Start report
"ultrasound   indication: rule out infiltrate/fluid  machine:  mindray   archived:  local   probe: 9-3 mhz  image quality: good   narrative: posterior , axillary and anterior acoustic windows interrogated bilaterally.  he has excess long b-lines an moth-eaten pleura and the lower right base only.  there is sparing of the eczema.  no consolidation.  no effusion impression: abnormal lung us  consistent with very mild viral pneumonitis.
"Mild"

Start report
"ultrasound  sutter medical center emergency department - sacramento    exam: lung ultrasound    indications: rule out pneumonia     probe: 10.5 mghz linear probe     image quality: good    procedure: using the linear array transducer i evaluated the bilateral anterolateral thorax in a systematic fashion revealing the presence of lung sliding and comet-tail

artifact at the interrogated interspaces. m-mode shows normal anatomy and function. findings: upper limit of normal b lines of bases of both lung fields. there is atelectasis noted on the left lung and a non-linear broncohogram definitive for infiltrate. negative for pleural effusions impression: no notable findings definitive for pneumonia. these images were archived and i independently interpreted the images at the bedside."
"Normal"

Start report
"ultrasound indication: rule out infiltrate/fluid machine: mindray archived: local probe: 10.5 mhz image quality: limited by child not staying still narrative: posterior , axillary and anterior acoustic windows interrogated bilaterally. normal a lines throughout. no excess b-lines, air bronchograms or effusion. no consolidation impression:normal lung us, upper limit of normal b lines "
"Normal"

Start report
"point of care lung ultrasound indication: rule out infiltrate/fluid machine: mindray archived: local probe: 10.5 mhz image quality: good narrative: posterior, axillar and anterior acoustic windows interrogated bilaterally. normal a-lines, no excess b-lines, no consolidation, no effusion impression: normal us "
"Normal"

Start report
"ultrasound indication: rule out infiltrate/fluid machine: mindray archived: local probe: 10.5 mhz image quality: good narrative: posterior , axillary and anterior acoustic windows interrogated bilaterally. normal a lines throughout. no excess b-lines, air bronchograms or effusion. no consolidation impression:normal lung us "
"Normal"

Start report
" point of care lung ultrasound indication: rule out infiltrate/fluid machine: mindray archived: local probe: 10.5 mhz image quality: very good narrative: posterior , axillar and anterior acoustic windows interrogated bilaterally. one area has borderline b-line. everything else is okay. impression: normal. t
"Very mild"

Start report
"ultrasound indication: rule out infiltrate/fluid machine: mindray archived: local probe: 10.5 mhz image quality: good narrative: posterior , axillary and anterior acoustic windows interrogated bilaterally. normal a lines throughout. no excess b-lines, air bronchograms or effusion. no consolidation. no pneumonia. impression:normal lung us "
"Normal"

Start report
"ultrasound indication: rule out infiltrate/fluid machine: mindray archived: local probe: 10.5 mhz image quality: good narrative: posterior , axillary and anterior acoustic windows interrogated bilaterally. normal a lines throughout. no excess b-lines, air bronchograms or effusion. no consolidation impression:normal lung us
"Normal"

Start report
"ultrasound indication: rule out infiltrate/fluid machine: mindray archived: local probe: 10.5 mhz image quality:

good narrative: posterior , axillary and anterior acoustic windows interrogated bilaterally. normal a lines throughout. no excess b-lines, air bronchograms or effusion. no consolidation impression:normal lung us abnormal lab results: no abnormal labs in the last 3 months. this document was transcribed by julianna c rojo, cms. signature & attestation: all medical record entries made by the scribe were at my direction. i have reviewed the chart and agree that the record accurately reflects my person"
"Normal"

Start report
" point of care lung ultrasound indication: rule out infiltrate/fluid machine: mindray archived: local probe: 10.5 mhz image quality:very good narrative: posterior , axillary and anterior acoustic windows interrogated bilaterally. he has crowded air bronchograms and a small area of consolidation between 0.5 and 1 cm depth in the right upper acoustic windows posteriorly and in the upper most portion of the right axillary window. does not have moth-eaten pleura or the coalescent b-lines typically seen with covid. impression: right upper lobe pneumonia "
"Mild to Moderate"

Start report
" point of care limited lung ultrasound indication: rule out infiltrate/fluid machine: mindray archived: local probe: 10.5 mhz image quality: good narrative: posterior , axillary and anterior acoustic windows interrogated bilaterally. he has moth-eaten pleura and occasional excess b-lines bilaterally. no effusion no consolidation. impression: very mild viral pneumonitis "
"Very mild"

Start report
"ultrasound indication: rule out infiltrate/fluid machine: mindray archived: local probe: 10.5 mhz image quality: good narrative: posterior , axillary and anterior acoustic windows interrogated bilaterally. normal a lines throughout. no excess b-lines, air bronchograms or effusion. no consolidation impression:normal lung us "
"Normal"

Start report
"ultrasound unremarkable. point of care lung ultrasound indication: rule out infiltrate/fluid machine: mindray archived: local probe: 10.5 mhz image quality: good narrative: posterior , axillary and anterior acoustic windows interrogated bilaterally. normal a lines throughout. no excess b-lines, air bronchograms or effusion. no consolidation impression: normal lung us "
"Normal"

Start report
" point of care lung ultrasound indication: rule out infiltrate/fluid machine: mindray archived: local probe: 10.5 mhz image quality: v good narrative: posterior acoustic windows interrogated bilaterally. normal a lines, few linear airbronchgram lul only c/w atelectasis., impression likely viral infection lul normal otherwise not covid records reviewed: nursing notes relevant past records "
"Very mild"

Start report
" point of care lung ultrasound indication: rule out infiltrate/fluid machine: mindray archived: local probe: 4.1mhz image quality: *** narrative: posterior , axillar and anterior acoustic windows interrogated bilaterally. increased breath sounds diffusely no effusion. normal a lines. normal b lines impression:*** *** "
"Normal"

Start report
" point of care lung ultrasound indication: rule out infiltrate/fluid machine: mindray archived: local probe: 10.5 mhz image quality: *** narrative: posterior , axillar and anterior acoustic windows interrogated bilaterally. normal a lines. normal b lines. no profusion. no consolidation. impression:normal us. "
"Normal"

Start report
"ultrasound indication: rule out infiltrate/fluid machine: mindray archived: local probe: 10.5 mhz image quality:good narrative: posterior , axillary and anterior acoustic windows interrogated bilaterally. normal a lines throughout. no excess b-lines, air bronchograms or effusion. no consolidation impression:normal lung us
"Normal"

Start report
" point of care lung ultrasound indication: rule out infiltrate/fluid machine: mindray archived: local probe: 10.5 mhz image quality: good narrative: posterior , axillar and anterior acoustic windows interrogated bilaterally. he does have borderline increased b lines bilaterally in the axilla right > left impression: possible mild pnemonitis "
"Mild"

Start report
"ultrasound indication: rule out infiltrate/fluid machine: mindray archived: local probe: 10.5 mhz image quality: good narrative: posterior , axillary and anterior acoustic windows interrogated bilaterally. normal a lines throughout. no excess b-lines, air bronchograms or effusion. no consolidation impression:normal lung us "
"Normal"

Start report
"ultrasound indication: rule out infiltrate/fluid machine: mindray archived: local probe: 10.5 mhz image quality: good narrative: posterior , axillary and anterior acoustic windows interrogated bilaterally. normal a lines throughout. no excess b-lines, air bronchograms or effusion. no consolidation impression:normal lung us "
"Normal"

Start report
"ultrasound. limited bedside poc ultrasound radiology report sutter medical center emergency department - sacramento exam: poc lung ultrasound indications: chest pain or shortness of breath views: 2-3 views (as below) probe: linear probe image quality: good procedure: using the linear array transducer i evaluated the bilateral anterolateral thorax in a systematic fashion revealing the presence of lung sliding and comet-tail artifact at the interrogated interspaces. m-mode shows normal anatomy and function. impression: negative limited lung ultrasound with no pneumothorax identif"
"Normal"

Start report
" point of care limited lung ultrasound indication: rule out infiltrate/fluid machine: mindray archived: local probe: 4.3 mhz image quality: good narrative: posterior , axillary and anterior acoustic windows interrogated bilaterally. he he has patchy areas of at least moderate severity excess b-lines and moth-eaten pleura. he also was fairly large areas that are not involved. no effusion. no consolidation. bilateral changes. worse on the left. impression: consistent with moderate severity bilateral covid pneumonitis "
"Moderate"

Start report
"point of care lung ultrasound  indication: rule out infiltrate/fluid  machine:  mindray   archived:  local   probe: 10.5 mhz  image quality: good  narrative: posterior , axillary and anterior acoustic windows interrogated bilaterally. *** impression:***    mostly normal some areas of excess air bronchobrows on right.   occasion irreg plurea and b lines mist blines of uper limit normal  both axxila upper llimt of b lines  impress  borderline us.     "
"Very mild"

Start report
"ultrasound completed.     limited bedside poc ultrasound radiology report  sutter medical center emergency department - sacramento    exam: poc lung ultrasound    indications: bronchiolitis     views: 2-3 views (as below)    probe: linear probe    image quality: good    procedure: using the linear array transducer i evaluated the bilateral anterolateral thorax in a systematic fashion revealing the presence of lung sliding and comet-tail artifact at the interrogated interspaces. m-mode shows normal anatomy and function.     impression: negative limited lung ultrasound
"Normal"

Start report
"ultrasound  indication: rule out infiltrate/fluid  machine:  mindray   archived:  local   probe: 10.5 mhz  image quality: good   narrative: posterior , axillary and anterior acoustic windows interrogated bilaterally. excess short and long b-lines to left base in posterior window and left axillary window. increase air bronchogram*** to left base. right lung has excess short b-lines anteriorly with right lung otherwise normal.   impression:left pneumonitis with possible right anterior pneumonitis. no effusi"
"Moderate"

Start report
"ultrasound  indication: rule out infiltrate/fluid  machine:  mindray   archived:  local   probe: 10.5 mhz  image quality: good   narrative: posterior , axillary and anterior acoustic windows interrogated bilaterally. excess short and long b-lines to left base in posterior window and left axillary window. increase air bronchograms in the  left base. right lung has excess short b-lines anteriorly with right lung otherwise normal.  no effusion. no consolidation   impression:left pneumonitis with possible right anterior pneumonitis.  "
"Moderate"

Start report
"  point of care limited  lung ultrasound  indication: rule out infiltrate/fluid  machine:  mindray   archived:  local   probe: 10.5 mhz  image quality: good   narrative: posterior , axillary and anterior acoustic windows interrogated bilaterally. abnormal long b lines in left upper posterior and axillary window. no effusion or consolidation.  impression: abnormal consistent with focal pneumonitis   "
"Very mild"

Start report
" bedside ultrasound of the posterior lungs showed no pleural irregularities, consolidations, or b lines consistent with covid findings   "          "
"Normal"

Start report

"ultrasound indication: rule out infiltrate/fluid machine: mindray archived: local probe: 10.5 mhz image quality: good narrative: posterior , axillary and anterior acoustic windows interrogated bilaterally. normal a lines throughout. no excess b-lines, air bronchograms or effusion. no consolidation impression:normal lung us "

Start report
"ultrasound indication: rule out infiltrate/fluid machine: mindray archived: local probe: 10.5 mhz image quality: good narrative: posterior , axillary and anterior acoustic windows interrogated bilaterally. normal a lines throughout. no consolidation or effusion. he had 2 areas in the upper anterior feels with moth-eaten pleura and long coalescent b-lines. impression:abormal lung us consistent with pleural scarring. in the context of a positive covid test likely represents lung involvement of the covid. "
"Very mild"

Start report
" point of care limited lung ultrasound indication: rule out infiltrate/fluid machine: mindray archived: local probe: 10.5 mhz image quality: good narrative: posterior , axillary and anterior acoustic windows interrogated bilaterally. impression: no evidence of pulmonary edema. reviewed and electronically signed by paul walsh md. medical decision making: differential diagnosis includes [but is not limited to] viral uri, influenza, bronchiolitis, "
"Normal"

Start report
" point of care limited lung ultrasound indication: rule out infiltrate/fluid machine: mindray archived: local probe: 10.5 mhz image quality: good narrative: posterior , axillary and anterior acoustic windows interrogated bilaterally. some air bronchograms on the right acoustic window, posterior. no effusion or consolidation. no b-lines. impression: consistent with mild bronchitis or atelectasis. "
"Mild"

Start report
" point of care limited lung ultrasound indication: rule out infiltrate/fluid machine: mindray archived: local probe: 10.5 mhz image quality: good narrative: posterior , axillary and anterior acoustic windows interrogated bilaterally. some air bronchograms on the right acoustic window, posterior. no effusion or consolidation. no b-lines. impression: consistent with mild bronchitis or atelectasis. "
"Mild"

Start report
"ultrasound indication: rule out infiltrate/fluid machine: mindray archived: local probe: 10.5 mhz image quality: good narrative: posterior , axillary and anterior acoustic windows interrogated bilaterally. normal a lines throughout. no excess b-lines, air bronchograms or effusion. no consolidation impression:normal lung us "
"Normal"

Start report
"ultrasound indication: rule out infiltrate/fluid machine: mindray archived: local probe: 10.5 mhz image quality: good narrative: posterior , axillary and anterior acoustic windows interrogated bilaterally. normal a lines throughout. no excess b-lines, air bronchograms or effusion. no consolidation impression:normal lung us "

"Normal"

Start report
" point of care lung ultrasound indication: rule out infiltrate/fluid machine: mindray archived: local probe: 10.5 mhz image quality: good narrative: posterior , axillar and anterior acoustic windows interrogated bilaterally. patchy areas of increased b lines. no effusion or consolidation impression:virla pneumonitis "
"Mild"

Start report
"ultrasound indication: rule out infiltrate/fluid machine: mindray archived: local probe: 10.5 mhz image quality: good narrative: posterior , axillary and anterior acoustic windows interrogated bilaterally. normal a lines throughout. no excess b-lines, air bronchograms or effusion. no consolidation impression:normal lung us "
"Normal"

Start report
"ultrasound indication: rule out infiltrate/fluid machine: mindray archived: local probe: 10.5 mhz image quality:good narrative: posterior , axillary and anterior acoustic windows interrogated bilaterally. normal a lines throughout. no excess b-lines, air bronchograms or effusion. no consolidation impression:normal lung us "
"Normal"

Start report
" point of care limited lung ultrasound indication: rule out infiltrate/fluid machine: mindray archived: local probe: 10.5 mhz image quality: good narrative: posterior , axillary and anterior acoustic windows interrogated bilaterally. some excess b lines and air bronchograms bilaterally, worse on the right. no effusion. impression:consistent with bronchiolitis "
"Mild"

Start report
"ultrasound indication: rule out infiltrate/fluid machine: mindray archived: local probe: 10.5 mhz image quality: good narrative: posterior , axillary and anterior acoustic windows interrogated bilaterally. normal a lines throughout. no excess b-lines, air bronchograms or effusion. no consolidation impression:normal lung us "
"Normal"

Start report
"point of care lung ultrasound indication: rule out infiltrate/fluid machine: mindray archived: local probe: 10.5 mhz image quality: good narrative: posterior , axillar and anterior acoustic windows interrogated bilaterally. impression:normal "
"Normal"

Start report
"ultrasound indication: rule out infiltrate/fluid machine: mindray archived: local probe: 10.5 mhz image quality: good narrative: posterior , axillar and anterior acoustic windows interrogated bilaterally. no excess b lines, effusion or consolidaton impression:normal "
"Normal"

Start report
"ultrasound indication: rule out infiltrate/fluid machine: mindray archived: local probe: 10.5 mhz image quality: good narrative: posterior , axillar and anterior acoustic windows interrogated bilaterally. no excess b lines, effusion or consolidaton impression:normal "
"Normal"

Start report
" point of care lung ultrasound indication: rule out infiltrate/fluid machine: mindray archived: local probe: 10.5 mhz image quality: good narrative: posterior , axillary and anterior acoustic windows interrogated bilaterally. normal a lines throughout. no excess b-lines, air bronchograms or effusion. no consolidation impression:normal lung us "
"Normal"

Start report
"ultrasound indication: rule out infiltrate/fluid machine: mindray archived: local probe: 10.5 mhz image quality: good narrative: posterior and axillar acoustic windows interrogated bilaterally. impression: diffuse a-lines, no evidence of b-lines, no effusion "
"Normal"

Start report
"ultrasound indication: rule out infiltrate/fluid machine: mindray archived: local probe: 10.5 mhz image quality: good narrative: posterior , axillary and anterior acoustic windows interrogated bilaterally. normal a lines throughout. no excess b-lines, air bronchograms or effusion. no consolidation impression:normal lung us "
"Normal"

Start report
"ultrasound radiology report sutter medical center emergency department - sacramento exam: poc limited ultrasound of *** indications: *** views: 2-3 views (as below) probe: linear image quality: excellent point of care limited lung ultrasound indication: rule out infiltrate/fluid machine: mindray archived: local probe: 9-3 mhz image quality: good narrative: posterior , axillary and anterior acoustic windows interrogated bilaterally. area of excess long b-lines in the right intrascapular window posteriorly. no effusion or consolidation. some moth eaten pleura bilaterally. impression: abnormal lung us reviewed and electronically signed b"
"Mild to Moderate"

Start report
"point of care lung ultrasound indication: rule out infiltrate/fluid machine: mindray archived: local probe: 10.5 mhz image quality: normal narrative: posterior acoustic windows interrogated bilaterally. impression: normal. normal a lines and b lines "
"Normal"

Start report

"  point of care limited  lung ultrasound  indication: rule out infiltrate/fluid  machine:  mindray   archived:  local probe: 10.5 mhz  image quality: good   narrative: posterior , axillary and anterior acoustic windows interrogated bilaterally. b lines upper limit of normal. there are a couple of air bronchograms.   impression: us more c/w non-covid infection.          "
"Very mild"

Start report
"  limited bedside poc ultrasound radiology report  sutter medical center emergency department - sacramento    exam: focus lung ultrasound, mindray     indications: cough     probe: 4.5mhz, 10.5mhz probe failed due to body habitus image quality: poor     archives: local     procedure: using the probe i evaluated the bilateral anterolateral thorax in a systematic fashion revealing the presence of lung sliding and comet-tail artifact at the interrogated interspaces. impression: borderline excess b line in right upper posterior acoustic window with thickening, subpleural less than 1cm. on the right there is an "
"Very mild"

Start report
"ultrasound  indication: rule out infiltrate/fluid  machine:  mindray   archived:  local   probe: 10.5 mhz  image quality: good   narrative: posterior , axillary and anterior acoustic windows interrogated bilaterally.  no effusion. area of air bronchograms on the left upper lobe less than 1 cm, bilateral long b lines in the bases and anterior lung fields. impression:  bilateral  long b lines in the bases and some airbronchogram? pneumonitis v bronchiolits v cld
"Moderate"

Start report
"ultrasound radiology report  sutter medical center emergency department - sacramento    exam: focus lung ultrasound indications: cough     probe: 10.5 mhz probe     image quality: good    procedure: ***     impression: upper limit of normal b lines on the right and left side    "      "
"Normal"

Start report
"ultrasound  indication: rule out infiltrate/fluid  machine:  mindray   archived:  local   probe: 10.5 mhz  image quality: ***  narrative: posterior , axillary and anterior acoustic windows interrogated bilaterally. normal a lines throughout. no excess b-lines, air bronchograms or effusion. no consolidation  impression:normal lung us
"Normal"

Start report
"ultrasound  indication: rule out infiltrate/fluid  machine:  mindray   archived:  local   probe: 10.5 mhz  image quality: good  narrative: posterior , axillary and anterior acoustic windows interrogated bilaterally. normal a lines throughout. no excess b-lines, air bronchograms or effusion. no consolidation  impression: normal lung us
"Normal"

Start report
"ultrasound  indication: rule out infiltrate/fluid  machine:  mindray   archived:  local   probe: 10.5 mhz  image quality:

adequate narrative: posterior , axillary and anterior acoustic windows interrogated bilaterally. impression: lung exam without abnormality, no focal infiltrate identified. medical decision making: differential diagnosis includes [but is not limited to] viral uri, asthma/copd exacerbation, pneumonia, pulmonary embolism among others in summary, this is a 5 month old female who presents with cough and rhinorrhea. evaluation in the ed most pertinent for reassuring pe a"
"Normal"

Start report
"ultrasound indication: rule out infiltrate/fluid machine: mindray archived: local probe: 10.5 mhz image quality: adequate narrative: posterior , axillary and anterior acoustic windows interrogated bilaterally. impression: lung exam without abnormality, no focal infiltrate identified.
"Normal"

Start report
"ultrasound indication: rule out infiltrate/fluid machine: mindray archived: local probe: 9-3 mhz image quality: good narrative: posterior , axillary and anterior acoustic windows interrogated bilaterally. she has an area of mild the pleura with excess long coalescent b-lines on the right posterior intrascapular windows. no effusion. no consolidation. impression: abnormal lung us focal unilateral viral pneumonitis
"Mild"

Start report
" point of care lung ultrasound indication: rule out infiltrate/fluid machine: mindray archived: local probe: 10.5 mhz image quality:fair narrative: posterior , axillar and anterior acoustic windows interrogated bilaterally. impression:excess b lines in all windows. based on bedside us, uncertain if it is viral pneumenitus or vs covid "
"Moderate"

Start report
" point of care limited lung ultrasound indication: rule out infiltrate/fluid machine: mindray archived: local probe: 10.5 mhz image quality: good narrative: posterior , axillary and anterior acoustic windows interrogated bilaterally. few b lines on the left base. no effusion/consolidation impression: mild focal pneumonitis left "
"Mild"

Start report
" point of care lung ultrasound indication: rule out infiltrate/fluid machine: mindray archived: local probe: 10.5 mhz image quality: good narrative: posterior , axillary, and anterior acoustic windows interrogated bilaterally. pleural thickening with excess non coalescent and coalescent lines in right upper anterior acoustic window. minimillay increased coalescent b lines posterior acoustic window. impression resolving multifocal viral pneumonitis
"Mild to Moderate"

Start report
"ultrasound indication: rule out infiltrate/fluid machine: mindray archived: local probe: 10.5 mhz image quality: *** narrative: posterior , axillary and anterior acoustic windows interrogated bilaterally. normal a lines throughout. no excess b-lines, air bronchograms or effusion. no consolidation impression:normal lung us "
"Normal"

Start report
" point of care lung ultrasound indication: rule out infiltrate/fluid machine: mindray archived: local probe: 10.5 mhz image quality: good narrative: posterior , axillar and anterior acoustic windows interrogated bilaterally. some increase b lines bilaterally, some exccess long b lines. mostly excess short b lines. no consolidation. no effusion. "
"Very mild"

Start report
" point of care lung ultrasound indication: rule out infiltrate/fluid machine: mindray archived: local probe: 10.5 mhz image quality: good narrative: posterior , axillar and anterior acoustic windows interrogated bilaterally. some increase b lines bilaterally, some excess long b lines. mostly excess short b lines. no consolidation. no effusion. impression:c/w covid or bronchiolitis . "
"Very mild"

Start report
" point of care lung ultrasound indication: rule out infiltrate/fluid machine: mindray archived: local probe: 10.5 mhz image quality: very good narrative: posterior and axillary acoustic windows interrogated bilaterally. he has an area pleural thickening and irregularity placed are bronchograms consistent with subcentimeter consolidation on the right. he has diffusely increased excessive coalescent b-lines and associated mop eaten pleura bilaterally in the intrascapular areas and in the axillary bilaterally. impression: small area of subcentimeter consolidation and associated bilateral diffuse pneumonitis worse on the right consistent with covid.
"Moderate"

Start report
" point of care lung ultrasound indication: rule out infiltrate/fluid machine: mindray archived: local probe: 10.5 mhz image quality: very good narrative: posterior examiner he and anterior acoustic windows interrogated bilaterally. he has upper limit of normal to slightly abnormal b lines with minimal pleural thickening only in the right upper most aspect of the posterior acoustic window. impression: fairly typical of what we would expect with rsv. does not appear to be i covid. "
"Very mild"

Start report
" point of care limited lung ultrasound indication: rule out infiltrate/fluid machine: mindray archived: local probe: 10.5 mhz image quality: good narrative: posterior , axillary and anterior acoustic windows interrogated bilaterally.minor excess short and some long b lines let base mostly no effusion no consolidation impression:mild covid pneumonitis "
"Mild"

Start report
"indicated) point of care lung ultrasound indication: rule out infiltrate/fluid machine: mindray archived: local probe: 10.5 mhz image quality: good narrative: posterior and axillary and anterior acoustic windows interrogated

bilaterally.increased, mostly short b-lines left axillary window. impression: cw mild covid medical decision ma"
"Mild"

Start report
"ultrasound indication: rule out infiltrate/fluid machine: mindray archived: local probe: 10.5 mhz image quality: *** narrative: posterior , axillary and anterior acoustic windows interrogated bilaterally. normal a lines throughout. no excess b-lines, air bronchograms or effusion. no consolidation impression:normal lung us
"Normal"

Start report
"ultrasound indication: rule out infiltrate/fluid machine: mindray archived: local probe: 10.5 mhz image quality: good narrative: posterior , axillary and anterior acoustic windows interrogated bilaterally. normal a lines throughout. no excess b-lines, air bronchograms or effusion. no consolidation impression:normal lung us "
"Normal"

Start report
"ultrasound indication: rule out infiltrate/fluid machine: mindray archived: local probe: 9-3 mhz image quality: good narrative: posterior , axillary and anterior acoustic windows interrogated bilaterally. the left posterior acoustic windows show increased air bronchograms diffusely. no excess b-lines. left axilla is normal. the right posterior acoustic window shows areas of excess long b-lines obliterating the a lines. in addition to this there was some hepatization and irregular air bronchograms. the consolidation is greater than 1 centimetre in depth. there is no effusion. impression: abnormal lung us consistent with right lower lobe pneumonia
"Mild to Moderate"

Start report
"point of care lung ultrasound indication: rule out infiltrate/fluid machine: mindray archived: local probe: 10.5 mhz image quality: excellent narrative: posterior , axillary and anterior acoustic windows interrogated bilaterally. excess b lines diffusely in both right and left intrascapular regions. impression: moderate pneumonitis consistent with covid "
"Moderate"

Start report
" limited bedside poc ultrasound radiology report sutter medical center emergency department - sacramento exam: poc lung ultrasound indications: irritability views: 2-3 views (as below) probe: linear probe image quality: good procedure: using the linear array transducer i evaluated the bilateral posterolateral thorax in a systematic fashion revealing the presence of lung sliding with pleural thickening and b-lines in multiple lung fields. impression: positive findings for covid-19. these images were archived and i independently interpreted the images at the bedside.
"Moderate"

Start report
"ultrasound radiology report sutter medical center emergency department - sacramento exam: poc lung ultrasound indications: cough views: 2-3 views (as below) probe: linear probe image quality: good procedure: using the linear array transducer i evaluated the bilateral anterolateral thorax in a systematic fashion revealing the presence of lung sliding and at the interrogated interspaces. m-mode shows normal anatomy and function. there are multiple mild b-lines in various lung fields bilaterally consistent with mild coronavirus pneumonitis impression: positive limited

lung ultrasound with no pneumothorax identified. exam consistent with mild covid pneumonitis.
"Mild"

Start report
" point of care lung ultrasound indication: rule out infiltrate/fluid machine: mindray archived: local probe: 10.5 mhz image quality: good narrative: posterior , axillar and anterior acoustic windows interrogated bilaterally. b lines on right axillar impression:concernign for pulmonary edema vs pneumonia, "
"Very mild"

Start report
"ed procedures: point of care lung ultrasound indication: rule out infiltrate/fluid machine: mindray archived: local probe: 10.5 mhz image quality: good narrative: posterior , axillar and anterior acoustic windows interrogated bilaterally. exces b lines on right axillary window impression:concernign for pulmonary edema vs pneumonia,
"Mild to Moderate"

Start report
" point of care lung ultrasound indication: rule out infiltrate/fluid machine: mindray archived: local probe: 10.5 mhz image quality: very good narrative: posterior , axillar and anterior acoustic windows interrogated bilaterally. diffuse excess b-line and pleural thickening bilaterally in all lung windows. impression:consistent with covid. "
"Moderate"

Start report
" point of care lung ultrasound indication: rule out infiltrate/fluid machine: mindray archived: local probe: 4.1mhz image quality: narrative: posterior , axillar and anterior acoustic windows interrogated bilaterally. she has excess long and short b lines and moth eaten pleura bilaterally impression: viral pneumonitis c/w covid "
"Mild to Moderate"

Start report
" point of care lung ultrasound indication: rule out infiltrate/fluid machine: mindray archived: local probe: 10.5 mhz image quality: good narrative: posterior , axillar and anterior acoustic windows interrogated bilaterally. normal a lines, no excess b lines, no effusion or consolidation impression:normal "
"Normal"

Start report
" point of care lung ultrasound indication: rule out infiltrate/fluid machine: mindray archived: local probe: 10.5 mhz image quality: *** narrative: posterior , axillar and anterior acoustic windows interrogated bilaterally. *** impression: lung upper limit has short b - lines in the left axilla. no effusion. no consolidation. "
"Normal"

Start report
"ultrasound indication: rule out infiltrate/fluid machine: mindray archived: local probe: 10.5 mhz image quality: very good narrative: posterior , axillar and anterior acoustic windows interrogated bilaterally. lung upper limit has short b - lines in the left axilla. no effusion. no consolidation. impression: normal "
"Normal"

Start report
"    point of care lung ultrasound  indication: rule out infiltrate/fluid  machine:  mindray   archived:  local   probe: 10.5 mhz  image quality: fair  narrative: posterior , axillar and anterior acoustic windows interrogated bilaterally. impression: normal   "
"Normal"

Start report
"  point of care lung ultrasound  indication: rule out infiltrate/fluid  machine:  mindray   archived:  local   probe: 10.5 mhz  image quality: ***  narrative: posterior, axillary and anterior acoustic windows interrogated bilaterally. impression:excess b-lines in two intercostal spaces, mild bronchiolitis or pneumonitis. no consolidation, no effusion. "
"Mild"

Start report
"point of care lung ultrasound  indication: rule out infiltrate/fluid  machine:  mindray   archived:  local   probe: 10.5 mhz  image quality: excellent  narrative: posterior , axillary and anterior acoustic windows interrogated bilaterally. impression:excess b lines mostly in the left. b lines slightly to the right. no effusion. no consolidation.                "
"Mild to Moderate"

Start report
"point of care lung ultrasound  indication: rule out infiltrate/fluid  machine:  mindray   archived:  local   probe: 10.5 mhz  image quality: good  narrative: posterior , axillar and anterior acoustic windows interrogated bilaterally. impression: normal us "      "
"Normal"

Start report
" point of care lung ultrasound  indication: rule out infiltrate/fluid  machine:  mindray   archived:  local   probe: 10.5 mhz  image quality: good  narrative: posterior , axillar and anterior acoustic windows interrogated bilaterally. impression:no b-lines to suggest pulmonary edema or covid-19 pna.      "
"Normal"

Start report
"ultrasound   indication: rule out infiltrate/fluid  machine:  mindray   archived:  local   probe: 10-3mhz  image quality: good   narrative: posterior , axillary and anterior acoustic windows interrogated bilaterally. normal a lines throughout. no excess b-lines, or effusion. no consolidation.  she does have some linear air bronchograms bilaterally consistent with atelectasis or early bronchiolitis.    impression:abnormal lung us however there are insufficient other findings to label bronchiolitis at this point and for now we will take a watchful waiting approach. "
"Very mild"

Start report
"ultrasound    indication: rule out infiltrate/fluid  machine:  mindray   archived:  local   probe: 9-3 mhz  image quality: good  narrative: posterior , axillary and anterior acoustic windows interrogated bilaterally. excess b lines in right axilla.

moth-eaten pleura bilaterally in the intrascapular windows.   impression: abnormal lung us consistent with mild covid pneumonitis  "
"Mild"
